# Long-term Prediction of Severe Hypoglycemia in Type 2 Diabetes Based on Multi-view Co-training

**DOI:** 10.1101/2023.08.08.23293518

**Authors:** Melih Agraz, Yixiang Deng, George Em Karniadakis, Christos Socrates Mantzoros

## Abstract

**Background:** Patients with type 2 diabetes mellitus (T2DM) who have severe hypoglycemia (SH) poses a considerable risk of long-term death, demanding urgent medical attention. Accurate prediction of SH remains challenging due to its multifactorial nature, contributed from factors such as medications, lifestyle choices, and metabolic measurements.

**Method:** In this study, we propose a systematic approach to improve the robustness and accuracy of SH predictions using machine learning models, guided by clinical feature selection. Our focus is on developing one-year SH prediction models using both semi-supervised learning and supervised learning algorithms. Utilizing the clinical trial, namely Action to Control Cardiovascular Risk in Diabetes, which involves electronic health records for over 10,000 individuals, we specifically investigate adults with T2DM who are at an increased risk of cardiovascular complications.

**Results:** Our results indicate that the application of a multi-view co-training method, incorporating the random forest algorithm, improves the specificity of SH prediction, while the same setup with Naive Bayes replacing random forest demonstrates better sensitivity. Our framework also provides interpretability of machine learning (XAI) models by identifying key predictors for hypoglycemia, including fast plasma glucose, hemoglobin A1c, general diabetes education, and NPH or L insulins.

**Conclusion:** By enhancing prediction accuracy and identifying crucial predictive features, our study contributes to advancing the understanding and management of hypoglycemia in this population.

## Background

Type 2 diabetes mellitus (T2DM) results from either reduced insulin production, insulin resistance, or both. T2DM outnumbers both gestational diabetes and type 1 diabetes mellitus (T1DM) in prevalence, accounting for nearly 90% of all diagnosed cases [1]. Hypoglycemia can potentially have a critical impact on morbidity and mortality risk in T2DM [2]. From a clinical perspective, hypoglycemia can be divided into two categories: mild hypoglycemia (MH) or severe hypoglycemia (SH), which is determined based on whether the patient experiences loss of consciousness or needs medical assistance [3]. One critical area of risk prediction is the estimation of SH in diabetes, as it is marked by the need for immediate medical assistance and it is believed to be a strong risk factor of long-term mortality [4, 5]. SH can lead to seizures, coma, and brain damage [6, 7, 8, 9], and sometimes it can be fatal. According to a recent analysis conducted by the Action to Control Cardiovascular Risk in Diabetes (ACCORD) study, the presence of SH with medical assistance was linked to a 50% higher likelihood of developing heart failure [10], and youngs with Type 1 diabetes mellitus may die from hypoglycemia in up to 10% of cases [11]. Therefore, predicting SH risks in advance is important to prevent future heart attacks and take precautions against the resulting impact. For this reason, the clinical motivation of the study is to help healthcare professionals evaluate SH risks, predict the SH events and take precautions, thus protecting patients from the side effects of SH events in the future.

Electronic health records (EHRs), which include comprehensive patient records such as demographics, laboratory results, diagnoses, and medical histories, are digitally maintained throughout the treatment or follow-up process [12, 13]. As large datasets become increasingly available and computing resources become more powerful, complex analyses that were previously not possible with statistical methods have been done using machine learning (ML) techniques, enabling more accurate and effective predictions, particularly in the medical field. The application of ML techniques in EHRs data has gained significant attention in recent years [14, 15, 16], with a specific focus on EHRs data for diabetes prediction and management [17, 18, 19]. Zheng et al. [18] suggested a semi-automated ML model to distinguish between individuals with and without T2DM. They wanted to raise recall rates while keeping false positive rates at a minimum. To accomplish this, they performed feature engineering and then trained various conventional ML models, k-nearest-neighbors, Naive Bayes (NB), decision tree, Random Forest (RF), support vector machine, and logistic regression, based on the selected features. By analyzing the EHRs of 300 patients between 2012-2014, this research has revealed a more precise and effective approach to identifying individuals with and without T2DM. Zou et al. [8] used ML techniques (decision tree, RF, and neural network) to predict diabetes based on data from Luzhou, China. They performed five-fold cross-validation and used principal component analysis and MRMR to reduce the dimensionality of the dataset. The authors found that the RF algorithm achieved the highest accuracy, 80.84%. Nguyen et al. [19] developed a hybrid system to predict the onset of diabetes by combining wide and deep learning models by using EHRs data. Their hybrid approach outperformed other models, achieving an accuracy level of 84.28%. This highlights the potential to use ML to improve diabetes risk prediction and management.

The ACCORD EHRs dataset used in this study is characterized by a considerable number of features. Using this large dataset, clinical researchers focusing on diabetes could conduct risk prediction studies to identify factors that contribute to SH. Recently, advancements in ML and statistical methods have opened new avenues for predicting diabetes progression more accurately, offering new opportunities for future research. Supervised learning (SL) is one of the most widely used ML sub-field in medical researches, where labeled data is used in the training process to make decisions or make predictions such as in classification problems in pediatrics [20], early diagnosis of cancer [21], identification of drug candidates [22], predicting 1-year cardiovascular events [23], and predicting blood glucose level in T2D [24]. While typical SL requires labeled data exclusively, it may not be always feasible in medical research due to limited availability or missing outcomes. Semi-supervised learning (SSL) methods address this limitation by incorporating unlabeled data i.e., SSL methods are another sub-field of ML that combines SL and unsupervised learning by using both labeled and unlabeled data together. In addition, it would be more reasonable to use SSL methods for clinical data, especially if the outputs contain unlabeled outcomes. The main purpose of SSL methods is to increase the performance of labeled data by using unlabeled observations or to increase the number of labels by producing pseudo labels. Due to these reasons, SSL models have been widely applied to medical studies by integrating labeled and unlabeled data, such as in cardiovascular risk prediction [25], diabetes disease diagnosis [26], breast cancer survival prediction [27] and early prediction of pregnancy-associated hypertension [28] and medical image analysis [29].

In this study, we use the ACCORD EHRs dataset and propose a multi-view co-training ML model as an effective SSL method for predicting SH events. The current study is the first to predict SH events, especially by proposing a multi-view co-training ML model. This choice is motivated by the unlabeled data available in the ACCORD dataset. In particular, we encountered the following two problems with the ACCORD dataset during our study: **i) Imbalanced data**. The dataset is highly imbalanced (see Fig. 2); while the imbalance rate is approximately 1:6.79 for the first year, this ratio increases to 1:120 by the end of the sixth year. **ii) Plurality of features**. The main objectives of this study are: (i) to develop a ML model for predicting long-term SH events in patients with T2DM, which will allow patients and medical doctors to take appropriate precautions; and (ii) to identify the most effective features for predicting long-term SH events. Specifically, we are proposing a new multi-view co-training model for the long-term prediction, that is, predicting the second year “(t+1)” SH events using variables in the first year (t), based on the ACCORD [30] dataset. By achieving these goals, we hope to contribute to the development of more effective methods for managing SH events in patients with T2DM. This research introduces one-year SH prediction models that use both SSL and SL algorithms, and Fig. 1 provides a comprehensive visual representation of our research methodology, highlighting the key components and steps involved in our study. In Fig. 1, Panel (A) presents the overall structure of the pipeline, highlighting the various stages involved in our study. Panel (B) focuses on the feature selection process, illustrating the implementation of feature selection algorithms such as medical selection criteria (selected features by medical doctors), LASSO, Boruta, and MRMR. We refer to medically selected features as “MD” in this study. Panels (C) and (D) present the step-by-step processes of the single-view and multi-view co-training ML algorithms, respectively.

**Figure 1:**
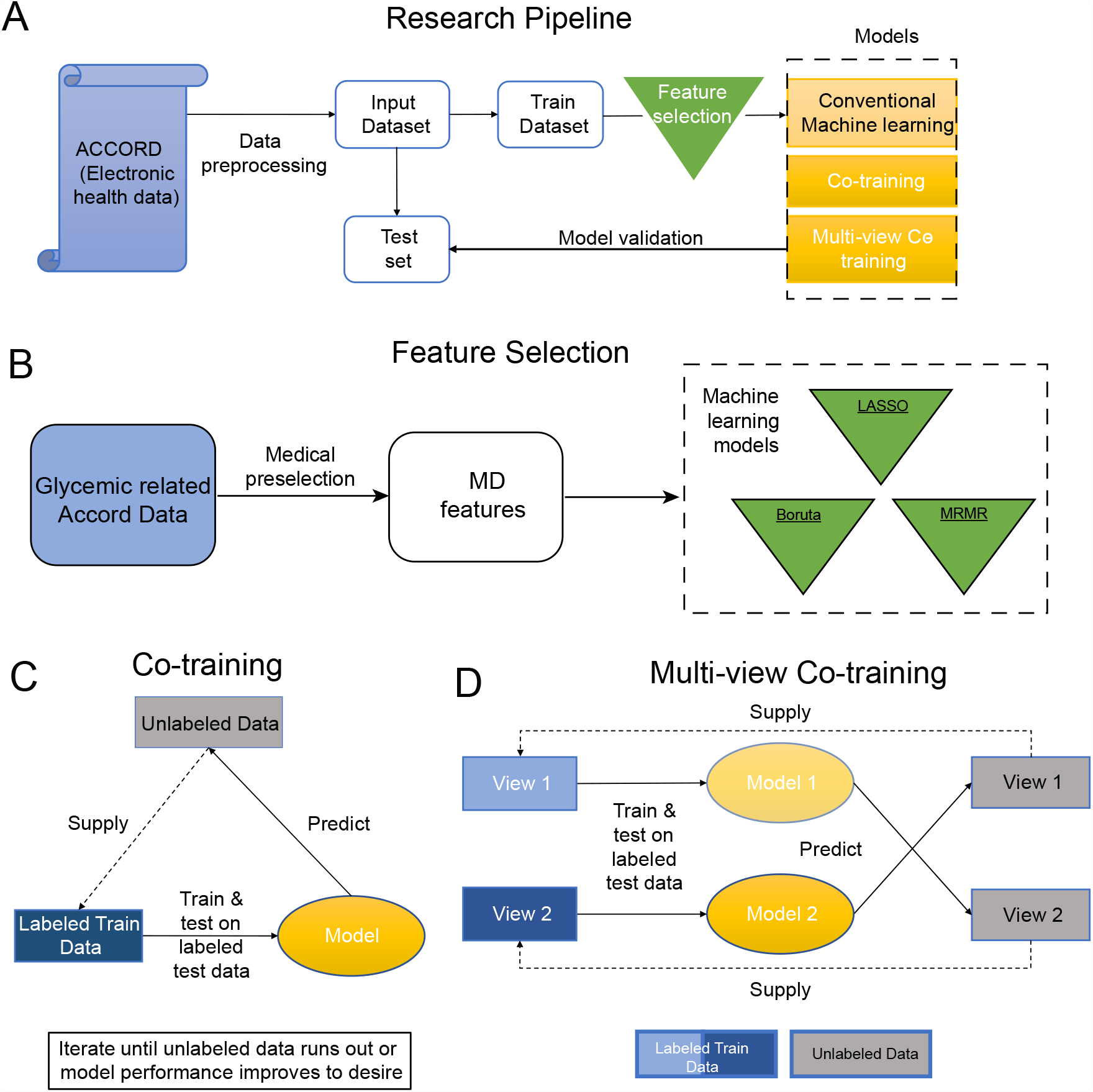
Research pipeline, feature selection methods and architecture of the models. (A)The overall structure of our research pipeline. **(B)** Diagram of the feature selection process. MD, LASSO, Boruta, and MRMR are feature selection algorithms. The output features of the LASSO, Boruta and MRMR feature selection methods are listed in Table 1. **(C)** Single-view co-training machine learning algorithm steps. **(D)** Multi-view co-training machine learning algorithm steps.

**Figure 2:**
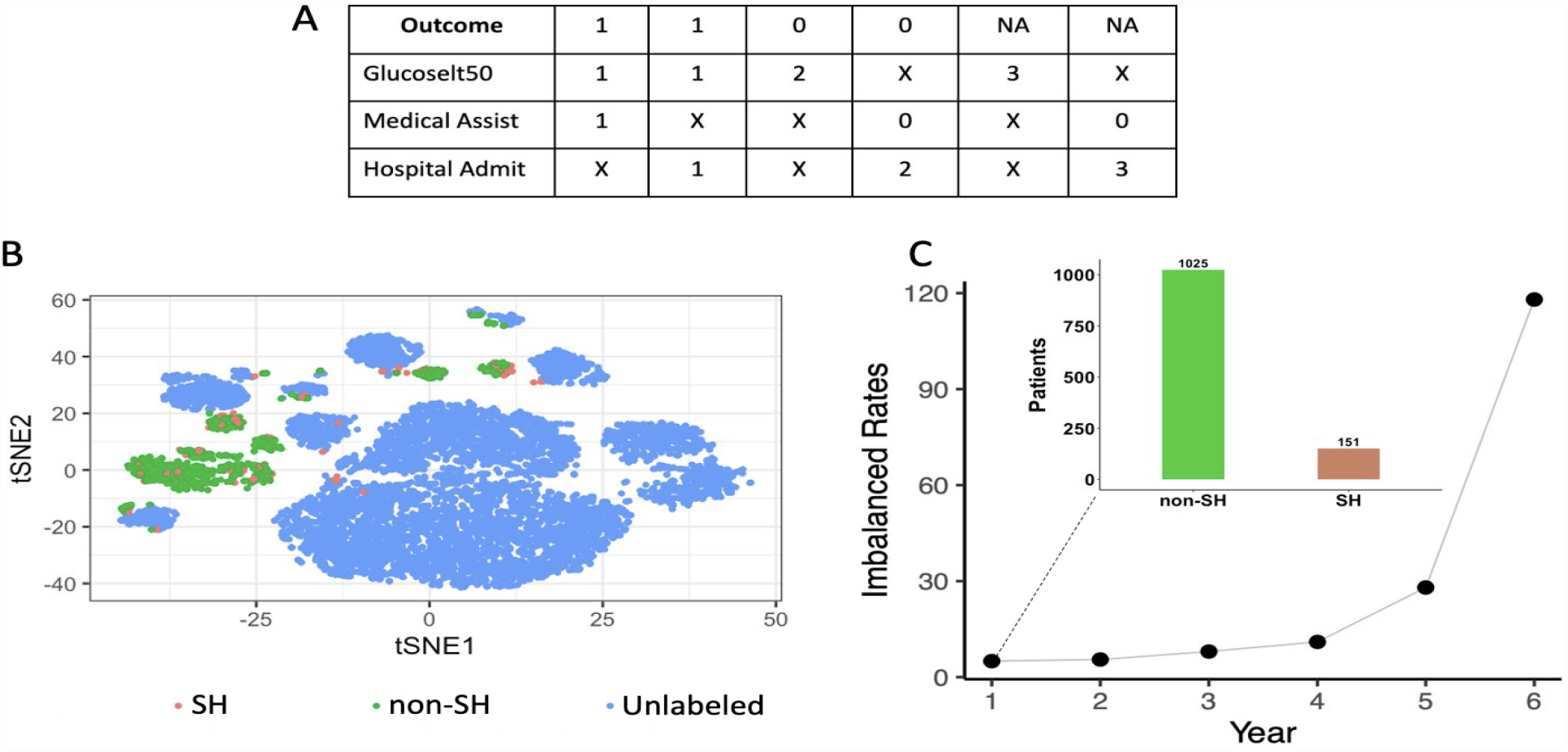
Visualization of the class of the dataset. **(A)** Related conditions for the dependent variable to be defined as SH. (1=Yes; 0 or 2=No; 3=Unknown; X= regardless of the value in the dataset).**(B)** Visualization of *t*-SNE of the high-dimensional dataset (17 dimensions) based on the ACCORD dataset colored by labeled (SH/non-SH) and unlabeled. Each dot represents a sample using MD. The labeled data is represented by green and red dots, corresponding to SH and non-SH events, respectively. Unlabeled data is represented by blue dots. **(C)** Hypoglycemia rates (non-SH/SH) of the dataset by year represented by solid circles. Class distribution of hypoglycemia events for the first year data represented by bar charts.

**Table 1:** ACCORD dataset MD features and the outputs after applying feature selection algorithms to them. The mean and standard deviation of the variables are calculated and included in the model. Bold features are effective features selected by the consensus and majority vote feature selection rule.

| MD Features | Description | Data Types | Selected by FS Algorithms |
| --- | --- | --- | --- |
| **HbA1c** | Glycated hemoglobin (%) | Continuous | LASSO, Boruta, MRMR |
| **FPG** | Fasting plasma glucose (mg/dL) | Continuous | LASSO, Boruta, MRMR |
| g1check | Average frequency of blood sugar check | Continuous | LASSO, Boruta |
| **g1diabed** | General diabetes education | Continuous | LASSO, Boruta, MRMR |
| g1nutrit | Nutrition education | Continuous | LASSO, Boruta |
| Sulfonylureas | Sulfonylureas | Categorical | LASSO |
| Meglitinides | Meglitinides | Categorical |  |
| **Nph/L Insulin** | NPH or L Insulins | Categorical | LASSO, Boruta, MRMR |
| Regular Insulins | Regular Insulins | Categorical |  |
| LA Insulin | Lispro or Aspart Insulins | Categorical |  |
| Othbol Insulin | Other Bolus Insulins | Categorical | LASSO, Boruta |
| Premixed Insulin | Premixed Insulins | Categorical | LASSO |

## Methods

### Missing data imputation

In this study, we applied two different methods, namely the last-observation-carried-forward (LOCF), for the time-series observations, and the median imputation, for the non-time-series observations, to handle missing data.

### Outcome

We determined the response variable of the ACCORD dataset according to Fig. 2A. According to Fig. 2A, “Glucoselt50” is assigned as a value of 1 if the blood glucose level is below 50 mg/dl, 2 if it is above 50 mg/dl, and 3 if no information is available (X: regardless of the value in the dataset). “Medical Assist” is assigned as 1, if medical assistance is required, 0 if not required. “Hospital Admit” is assigned as a value of 1, if the hospital admission is required, 2 if it is not required, and 3 if there is no information. Finally, the outcome is assigned as 1, 0, or NA based on the information provided by the patients. Patients who provided the following information were assigned as 1, i.e., as SH event; patients with a Blood Glucose level below 50 and either requiring Medical Assistance or Hospital Admission. For patients who provided the following information is assigned as 0, i.e., as non-SH; Blood Glucose is higher than 50. In addition, no medical assistance is required, and hospital admission is not needed assigned as value 0, indicating it is a non-SH event. Patients for whom we could not obtain information were assigned as NA, i.e. as missing-SH event; if the Blood Glucose information is unknown or Medical Assistance is not required and unknown Hospital Admission.

### Unlabeled dataset

The ACCORD dataset contains 9068 unlabeled and 1176 labeled data. First, we started working with the labeled dataset, but we could not obtain significant results, and subsequently we decided to include the unlabeled data in our analysis.

### Predictors

First, we created all 118 candidate risk features listed in supplementary material (SM) Table 3. After that, as represented in Fig. 1B, we selected the top-12 risk factors among the 118 SH relevant measurements from the ACCORD dataset. The MD estimators of the ACCORD dataset were chosen as follows: **HbA1c, FPG, g1check, g1diabed, g1nutrit, Sulfonylurea, Meglitinide, NPHL Insulin, Reg Insulin, La Insulin, Othbol Insulin, Premix Insulin**. As some of the features are longitudinal, we computed the mean and standard deviations of the observations and this process resulted in the following 17 variables: hba1c mean, hba1c std, fpg mean, fpg std, g1check mean, g1check std, g1diabed mean, g1diabed std, g1nutrit mean, g1nutrit std, sulfonylurea mean, meglitinide mean, nphl mean, reg insulin mean, la insulin mean, othbol insulin mean, premix insulin mean (See the SM Table 9).

### Views

In our study, we propose a multi-view co-training ML model as SSL. To begin the analysis, we first started with three different views. These are, glycemic variables (View 1): FPG, HBA1C; glycemic management and medications (View 2): g1check, g1diabed, g1nutrit, sulfonylurea, meglitinide, NPHL insulin, reg insulin, la insulin, othbol insulin, and premix insulin; (View 3): years of diabetes, live alone, education level, body mass index (BMI), participant waist circumference (cm), race, age, and gender. We examined and compared these three views and ranked View 1 and View 2 as more effective. Therefore, we generated two views for classification, glycemic variables based (View 1) and glycemic management and medications based (View 2).

### Time horizon

The ACCORD dataset participants were followed for approximately 4 to 8 years [30]. We decided to work on the second-year prediction, because the least imbalanced rate is seen for the first year.

### Feature selection and model validation

The feature selection algorithm is a method that helps to identify the most relevant variables from the input data and reduces it to a lower-dimensional dataset. Feature selection methods for classification tasks can be categorized into two groups [31]: expert knowledge-based feature selection methods, and automatic feature selection methods such as filter, wrapper, and embedded feature selection algorithms. In particular, we utilized the Boruta, MRMR, and LASSO methods as automatic feature selection algorithms. Furthermore, we not only evaluate the individual performances of these three feature selection methods but also consider the features that are selected by all of them as effective features. We incorporated a technique into our analysis, namely the “**consensus and majority vote feature selection**” rule [32], where the feature is considered an important feature if it is selected by all of the base feature selection methods in agreement. We have provided an explanation of the feature selection algorithms in the Feature Selection Methods subsection of the SM.

## Machine learning models

### Classification pipeline

All classifiers in this study are created using the caret package [33] in R programming language 4.1.3. We start by employing conventional ML algorithms to process the entire labeled dataset. Across the entire study, the dataset is split into two sets (see Fig. 1A): A 20% sample of the data is used to test the classifier’s performance, while the remaining 80% is used to train the classifier. We built models using the training dataset and tested their performance with 5-fold cross-validation. We first assessed the performance of several distinct classifiers, including LR, XGBoost, NB, SVM, and RF on the ACCORD dataset by calculating the classification accuracy. Then, we further evaluated the performance of single-view co-training and multi-view co-training models with NB and RF models.

1. **Naive Bayes classifier** A classification algorithm that operates on Bayes’ theorem and involves probabilities.
2. **Random forest** An ensemble classification or regression method that uses the decision tree algorithms[34].

### Single-view co-training model

It is obvious in Fig. 1C that labeled data is split as train and test set, and the model is initially trained (Step 1). Afterward, the trained model is used to estimate the unlabeled data (Step 2), and the most confident pseudo-labels are selected by the probability Θ higher than 0.90. In the next step (Step 3), the pseudo-labeled data and labeled data are concatenated. The model then makes predictions on unseen test data (Step 4), and finally, the results are evaluated (Step 5). Step 1 and Step 5 are repeated until new unlabeled data can no longer be added. We also tested the heterogeneity of the data by applying the cross-validation method.

### Multi-view co-training model

Blum [35] introduced the co-training algorithm, which is a semi-supervised learning algorithm, and numerious studies have been conducted on this topic [36, 37, 38]. The multi-view co-training method utilizes both views in tandem to supplement a much smaller number of labeled examples with unlabeled data. Blum [35] first defined the labeled (*L*), unlabeled dataset (*U*) and unlabeled pools (*U*′) (created for each View 1 and View 2), and set the number of iteration *k*, then divided the input space *X* = *X*_1_ *× X*_2_, so that *X*_1_ and *X*_2_ corresponding to two distinct sufficient and redundant views (View1 and View2) of the *X*, and they trained each view from the labeled data (*L*) by *h*_1_ and *h*_2_ classifiers. Then, the co-training method allows *h*_1_ to label *p* positive and *n* negative most confident labels from the unlabeled (*U*′) set (for View 2) as a pseudo label and again *h*_2_ to label *p* positive and *n* negative most confident labels from the unlabeled (*U*′) set (for View 1), so this prevents from over-training. Finally, the algorithm adds these confident labels to *L* and deletes the selected confidence labels from *U*′ (see Fig. 1D). Thus, the multi-view co-training method allows learning from both a few labeled and unlabeled data.

### Combining the views

Instead of performing classification, multi-view co-training is typically utilized to generate larger labeled data. In order to use it as a classification tool, it is necessary to combine the views generated at the end of the iteration within the multi-view co-training. There are many methods to combine the views, but we prefer the naive AND and OR rule to combine final predictions coming from View 1 and View 2. The **AND** rule assigns the result as 1, if the results from the *i*th observation of both views are 1. The **OR** rule assigns the result as 1, if only one of the results from the *i*th observation from both views is 1.

## Results

### Study population

Data from 10,251 enrolled participants with clinical diagnoses of T2DM were collected through the ACCORD study. The study’s participants were mostly middle-aged and elderly patients ranging from 40 to 82, with an average of 62.2 years and an average diabetes duration of 10 years. Of the total participants, the majority were white (64.8%) and male (61.4%). In our study, after performing missing data imputation, we proceeded with the analysis using a total of 10,244 observations. Table 4 in SM displays the mean ± standard deviations and percentages (%) for the selected variables in the ACCORD dataset. Abbreviations used in the study are listed in Table 5.

### Data quality checking

Upon completing the labeling process based on Fig. 2A, we obtained a total of 1,176 labeled data points, consisting of 151 cases of SH and 1,025 non-SH cases, along with 9,068 unlabeled data points. Fig. 2B is a projection of the data obtained using the t-distributed stochastic neighbor embedding (*t*-SNE) method [39], as we revealed by the *t*-SNE, there is no clear separation based on the target classes of SH non-SH in labeled data. For this reason, we thought that we could obtain meaningful information from the unlabeled data. In addition, the SH rates of these patients for 6 years are shown in Fig. 2C for each year.

### Conventional machine learning results

We first analyzed and compared various feature selection algorithms on a labeled dataset. First, to prevent overfitting and informed by clinical knowledge on SH, we performed our analysis using a total of 17 features. Consequently, we performed feature selection with four different algorithms, MD, LASSO [40], Boruta [41] and MRMR [42, 43], as shown in Fig. 1B. Following the feature selection process, we compared the performance of representative ML algorithms. In the testing data, as seen in Table 2, the NB model with MRMR feature selection shows the best performance among all classifiers with specificity and accuracy of 0.740 and 0.696, respectively. Also, the NB model with LASSO feature selection demonstrates the highest performance for NPV and sensitivity of 0.942, and 0.986, respectively. Lastly, the RF model with MD achieves the highest performance with a PPV of 0.194 and an F1-score of 0.303. Additionally, the 5-fold AUC-ROC curves of the conventional ML algorithms can be viewed in SM Fig. 8. We have provided an explanation of the formula for performance measures in the Performance Measures subsection of the SM.

**Table 2:** Comparison of results from conventional models using MD, LASSO, Boruta, and MRMR feature selection algorithms. NPV: Negative predictive value; PPV: Positive predictive value; Spec: Specificity; Sens: Sensitivity; Acc: Accuracy; F1: F1-score. In terms of predictive performance, the NB model outperforms all other models.

| MD Feature Selection |  |  |  |  |  |  |
| --- | --- | --- | --- | --- | --- | --- |
| Algorithms | NPV | PPV | Spec | Sens | Acc | F1 |
| LR | 0.913 | 0.184 | 0.591 | 0.612 | 0.594 | 0.280 |
| XGBoost | 0.907 | 0.170 | 0.560 | 0.603 | 0.565 | 0.263 |
| NB | 0.924 | 0.131 | 0.038 | 0.979 | 0.159 | 0.230 |
| SVM | 0.912 | 0.188 | 0.617 | 0.597 | 0.614 | 0.285 |
| RF | 0.927 | **0.194** | 0.572 | 0.697 | 0.588 | **0.303** |

| LASSO Feature Selection |  |  |  |  |  |  |
| --- | --- | --- | --- | --- | --- | --- |
| Algorithms | NPV | PPV | Spec | Sens | Acc | F1 |
| LR | 0.906 | 0.180 | 0.611 | 0.567 | 0.605 | 0.271 |
| XGBoost | 0.909 | 0.172 | 0.559 | 0.617 | 0.566 | 0.267 |
| NB | **0.942** | 0.130 | 0.026 | **0.986** | 0.150 | 0.229 |
| SVM | 0.911 | 0.192 | 0.629 | 0.583 | 0.622 | 0.286 |
| RF | 0.924 | 0.191 | 0.578 | 0.681 | 0.590 | 0.297 |

| Boruta Feature Selection |  |  |  |  |  |  |
| --- | --- | --- | --- | --- | --- | --- |
| Algorithms | NPV | PPV | Spec | Sens | Acc | F1 |
| LR | 0.909 | 0.181 | 0.597 | 0.594 | 0.597 | 0.276 |
| XGBoost | 0.908 | 0.168 | 0.543 | 0.627 | 0.553 | 0.264 |
| NB | 0.938 | 0.130 | 0.027 | **0.986** | 0.150 | 0.230 |
| SVM | 0.912 | 0.187 | 0.608 | 0.601 | 0.607 | 0.284 |
| RF | 0.927 | 0.192 | 0.568 | 0.699 | 0.584 | 0.301 |

| MRMR Feature Selection |  |  |  |  |  |  |
| --- | --- | --- | --- | --- | --- | --- |
| Algorithms | NPV | PPV | Spec | Sens | Acc | F1 |
| LR | 0.910 | 0.184 | 0.623 | 0.574 | 0.617 | 0.277 |
| XGBoost | 0.906 | 0.166 | 0.552 | 0.610 | 0.559 | 0.259 |
| NB | 0.894 | 0.184 | **0.740** | 0.407 | **0.696** | 0.248 |
| SVM | 0.910 | 0.181 | 0.606 | 0.586 | 0.604 | 0.276 |
| RF | 0.917 | 0.180 | 0.559 | 0.657 | 0.571 | 0.282 |

**Table 3:** Comparing multi-view co-training model results on MD and MRMR selected data with an under-sampling imbalanced solution. Results of the MD data and MRMR data. Percentage=(ViewX.last-ViewX.1st) *×* 100/ViewX.1st. Both NB and RF achieve great improvements using these two selected features.

| NB |  |  |  |  |  |  |  |  |  |  |  |  |
| --- | --- | --- | --- | --- | --- | --- | --- | --- | --- | --- | --- | --- |
|  | MD |  |  |  |  |  | MRMR |  |  |  |  |  |
|  | NPV | PPV | Spec | Sens | Acc | F1 | NPV | PPV | Spec | Sens | Acc | F1 |
| View1.1st | 0.883 | 0.168 | 0.775 | 0.311 | 0.714 | 0.217 | 0.887 | 0.192 | 0.804 | 0.314 | 0.740 | 0.233 |
| View1.last | 0.884 | 0.204 | 0.867 | 0.230 | 0.785 | 0.212 | 0.883 | 0.213 | 0.874 | 0.224 | 0.790 | 0.214 |
| Percentage | **0.075%** | **21.238%** | **11.971%** | -25.943% | **9.878%** | -2.180% | -0.457% | **10.870%** | **8.674%** | -28.643% | **6.785%** | -8.149% |
| View2.1st | 0.905 | 0.157 | 0.485 | 0.649 | 0.506 | 0.252 | 0.910 | 0.172 | 0.570 | 0.609 | 0.575 | 0.267 |
| View2.last | 0.876 | 0.149 | 0.842 | 0.190 | 0.758 | 0.155 | 0.876 | 0.176 | 0.876 | 0.155 | 0.784 | 0.155 |
| Percentage | -3.174% | -4.844% | **73.755%** | -70.640% | **49.759%** | -38.270% | -3.720% | **2.066%** | **53.583%** | -74.495% | **36.396%** | -41.921% |

| RF |  |  |  |  |  |  |  |  |  |  |  |  |
| --- | --- | --- | --- | --- | --- | --- | --- | --- | --- | --- | --- | --- |
|  | MD |  |  |  |  |  | MRMR |  |  |  |  |  |
|  | NPV | PPV | Spec | Sens | Acc | F1 | NPV | PPV | Spec | Sens | Acc | F1 |
| View1.1st | 0.898 | 0.162 | 0.564 | 0.568 | 0.564 | 0.251 | 0.897 | 0.157 | 0.544 | 0.579 | 0.548 | 0.246 |
| View1.last | 0.891 | 0.183 | 0.757 | 0.368 | 0.708 | 0.244 | 0.884 | 0.161 | 0.747 | 0.333 | 0.693 | 0.216 |
| Percentage | -0.789% | **12.805%** | **34.099%** | -35.115% | **25.491%** | -2.853% | -1.502% | **2.817%** | **37.150%** | -42.596% | **26.345%** | -12.420% |
| View2.1st | 0.906 | 0.163 | 0.528 | 0.624 | 0.541 | 0.258 | 0.931 | 0.161 | 0.393 | 0.785 | 0.444 | 0.266 |
| View2.last | 0.886 | 0.177 | 0.789 | 0.308 | 0.728 | 0.224 | 0.874 | 0.221 | 0.961 | 0.065 | 0.845 | 0.136 |
| Percentage | -2.273% | **8.279%** | **49.543%** | -50.626% | **34.592%** | -13.252% | -6.130% | **37.319%** | **144.732%** | -91.745% | **90.416%** | -49.023% |

**Table 4:** Performance comparison of the conventional, single-view co-training and multi-view co-training ML model results. Multi-view co-training performs the best, but only the conventional NB LASSO meets the clinical criteria of high sensitivity.

| Method | NPV | PPV | Spec | Sens | Acc | F1 |
| --- | --- | --- | --- | --- | --- | --- |
| Conventional NB LASSO | **0.942** | 0.130 | 0.026 | **0.986** | 0.150 | 0.229 |
| Conventional RF MD | 0.927 | 0.194 | 0.572 | 0.697 | 0.588 | **0.303** |
| Single-view co-training RF MD | 0.924 | 0.186 | 0.561 | 0.689 | 0.577 | 0.293 |
| Multi-view co-training RF MD-OR Rule | 0.906 | 0.173 | 0.584 | 0.589 | 0.585 | 0.267 |
| Multi-view co-training RF MRMR-AND Rule | 0.873 | **0.300** | **0.993** | 0.020 | **0.868** | 0.037 |

**Table 5:**
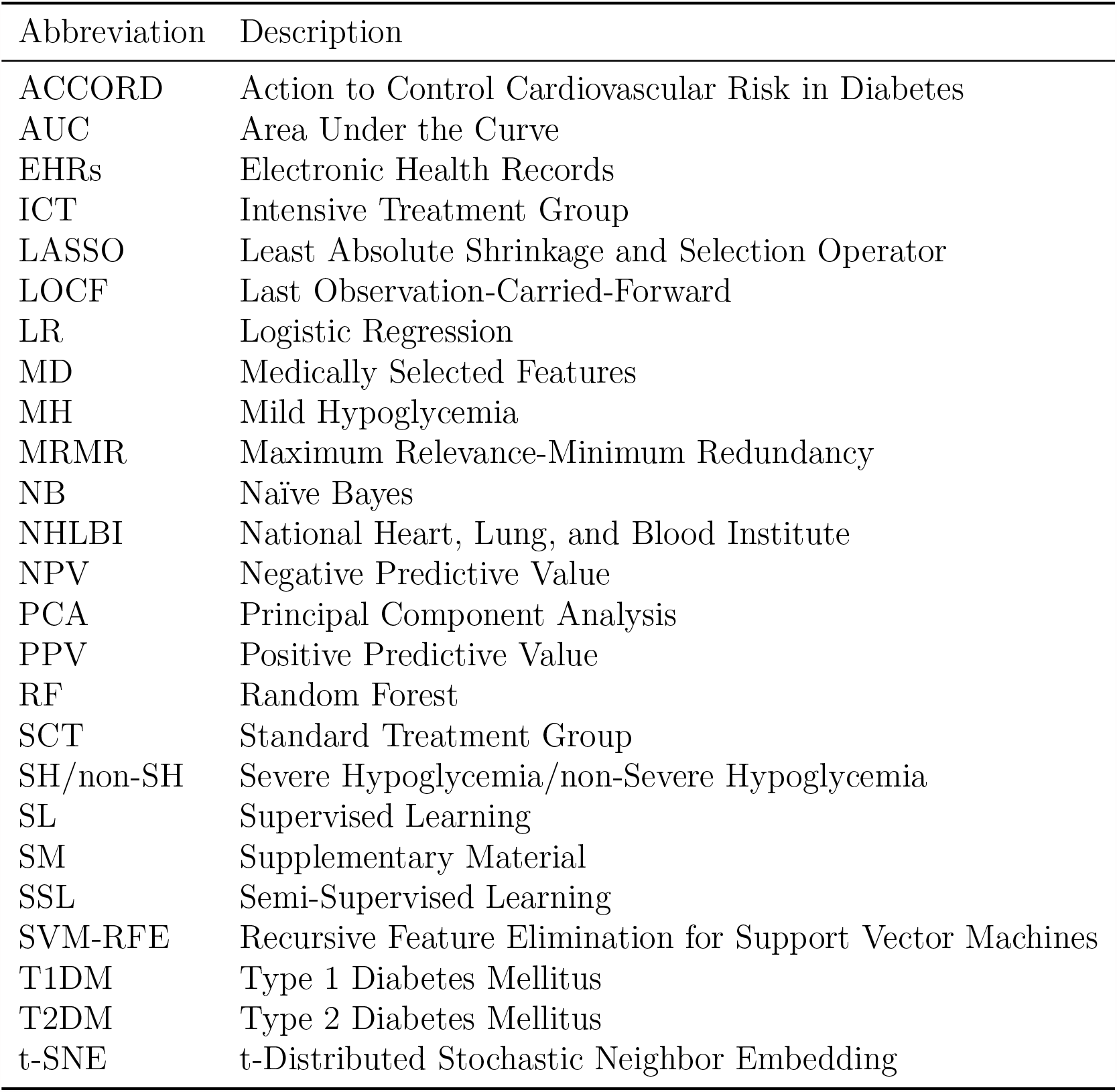
Abbreviations used in the study.

### Results based on the single-view co-training model

Despite the success of conventional models in demonstrating good performance in NPV, these models have shown limitations in other metrics that are crucial for practical implementation. Inspired by SSL, where integrating unlabeled datasets increase the performance of ML algorithms, we aim to incorporate a multitude of unlabeled observations in ACCORD data that remained to be explored. Due to limitations of space, we include the results for the single-view co-training model in the SM Table 1. As the single-view co-training method was not effective enough in improving conventional ML results, we switched to a different and more effective SSL method, the multi-view co-training method, as we discuss next. In addition, the confusion matrix of the single-view co-training results is listed in SM Fig. 6 and and mean accuracy metrics of test results for each iteration in single-view co-training for NB using MD features can be seen in SM Fig. 10.

### Results based on the multi-view co-training model

We fixed the number of iterations at 30, as experimented by Blum [35]. However, we executed different ratios of positive and negative pseudo-labels for selection, and we list the 5 negative/1 positive selection results in Table 3. Table 3 shows the accuracy measures for the first and the last iterations, as well as the percentage of gain or loss for each model and view. We highlight the positive contributions for five selected negative and one positive pseudo confident label. Table 3 indicates that in View 2, the highest contribution rates are observed for specificity (144.732%) and accuracy (90.416%) in the RF MRMR feature selection model. Additionally, performance curves of the accuracy, NPV, and AUC-ROC results are presented in SM Fig. 3. Moreover, we provide the confusion matrix of the multi-view co-training model results in SM Fig. 7, while in SM Fig. 9 displays the 5-fold AUC-ROC curves of the multi-view co-training algorithms.

### Combining the views of multi-view co-training model

Up to now, we have assessed the performance of each view created through the multi-view co-training model. Subsequently, we interpret the results of combining the information obtained from these views. Multi-view co-training is often used to increase the number of labeled data, but to employ multi-view co-training as a classification method, we need to combine the outputs. We employ a naive approach where the results are combined using both AND and OR rules. AND and OR rule results can be seen in SM Table 2 and the comparison of these combined results with other findings is presented in Table 4, showing the top-performing outcomes. We list the 5 negative/1 positive selection results in SM Fig. 1 while the other results (3 negative and 1 positive, and 7 negative and 1 positive) can be found in the SM Tables 5-8 and in SM Fig. 4-5.

Accordingly, we note that the AND rule produces better results for specificity, accuracy, and PPV measures, with values of 0.993, 0.868, and 0.300, respectively. On the other hand, the OR rule outperforms in terms of NPV, sensitivity, and F1, with values of 0.906, 0.589, and 0.267, respectively. (see more details in SM).

### Summarizing the best models

Finally, we can compare the top-performing results from conventional, single-view co-training, and multi-view co-training ML algorithms in Table 4. It is seen in Table 4 that the conventional models are only successful in predicting the majority class (non-SH) events, with NPV at 0.942 and sensitivity at 0.986, respectively. On the other hand, the multi-view models demonstrate a higher success rate in predicting SH events, achieving the best result for PPV at 0.300, specificity at 0.993, and accuracy at 0.868, respectively.

## Discussion

In this study, ML models were developed to predict the occurrence of SH events and identify the effective features that contribute to such predictions for patients with T2DM in the ACCORD data. The key findings of the study are: (1) top-12 features were selected by expert knowledge-based selection for predicting SH events; (2) one-year SH prediction models were developed using both SSL with a multi-view co-training method and SL with RF or NB models; and (3) the most effective features were proposed based on both expert knowledge-based selection and automatic feature selection methods for predicting SH events. (4) a shiny app [44] was designed to facilitate further analysis by researchers using the proposed multi-view co-training ML method.

The findings indicate that the suggested multi-view co-training approach exhibits superior performance in attaining elevated levels of specificity, positive predictive value (PPV), and overall accuracy. However, conventional ML algorithms surpass it in terms of sensitivity, negative predictive value (NPV), and F1 measures when predicting SH. Throughout this study, we employed different feature selection techniques prior to constructing the predictive model. Specifically, we explored the most suitable methods from both automatic and expert knowledge-based feature selection approaches. Specifically, we applied Boruta, MRMR, and LASSO method for automatic feature selection algorithms, while the expert knowledge-based method used in the study was referred as the MD method. Nevertheless, we achieved better results in predicting SH when we used automatic feature selection methods rather than merely MD method. Among them, we learned that the MRMR feature selection algorithm was the most effective at identifying the optimal features for the multi-view co-training model. In addition, we identified the effective features for the one-year SH prediction that are the common selection of these four features using the “the consensus and majority vote feature selection” rule [32]; **fast plasma glucose (FPG), hemoglobin A1c (HbA1c), general diabetes education (other than nutrition)(g1diabed) and NPH or L Insulins (NPHL insulin)**. Our framework also provides explainable artificial intelligence (XAI) [45] by identifying key predictors for hypoglycemia. Notably, HbA1c, FPG and NPH or L Insulins are beyond the control of patients and medical doctors, but general diabetes education can be controlled, and the development of effective self-management skills in patients has the potential to lower the incidence of hypoglycemia and increase knowledge regarding hypoglycemia [46]. Additionally, there is a risk between HbA1c [47, 48, 49] or FPG [47, 48] and higher risk of hypoglycemia. Finally, the most promising results are obtained by implementing the RF multi-view co-training method with the MRMR feature selection using the AND rule; NPV: 0.873, PPV: 0.300, specificity: 0.993, and accuracy: 0.868 and conventional NB model with LASSO feature selection; NPV:0.942, and sensitivity: 0.986. Multiple ML approaches were considered in this study, in order to determine the **“best”** modeling technique. However, it is crucial to know that there is not just one ideal model. For example, if the goal is to avoid SH, then “best” might be the model with the highest sensitivity [50], or high specificity would be “best” if the goal is to avoid unnecessary treatments or interventions. Therefore, we recommend the conventional NB LASSO to avoid SH, and the multi-view co-training RF MRMR-AND rule to avoid unnecessary intervention.

## Conclusion

We proposed ML methods in predicting SH events for T2DM patients and systematically examined our approaches using a seven-year clinical trial, namely ACCORD. We used SSL, specifically a novel multi-view co-training method, with SL methods, such as RF and NB, to improve accuracy. The study identified four key features that were highly effective in SH prediction: fast plasma glucose (FPG), hemoglobin A1c (HbA1c), general diabetes education (other than nutrition) (g1diabed), and NPH or L Insulins (NPHL insulin). For this reason, this study offers XAI, revealing the factors that influence the predictions. Our results suggest that the multi-view co-training approach can significantly improve the PPV, specificity and accuracy of SH event predictions. Our proposed multi-view co-training method can use data with missing labels, and this could benefit the T2DM patient population by improving SH event prediction accuracy and ultimately leading to more personalized treatment plans and overall improved patient health management. Additionally, using the proposed features can help clinicians to make better decisions. Finally we created a shiny app [44] for the proposed multi-view co-training method hosted on https://datascicence.shinyapps.io/MultiViewCoTraining/. In this app, the user can select the views and run multi-view co-training algorithm on naive bases (nb) and random forest (rf).

## Data Availability

The Action to Control Cardiovascular Risk in Diabetes (ACCORD) data is available upon request from the National Heart, Lung, and Blood Institute (NHLBI).

## Code availability

All codes are available on GitHub at http://github.com/melihagraz/XX.

## Declarations

### Ethics approval and consent to participate

The ethics committee of Beth Israel Deaconess Medical Center determined that this study was exempt re: review and approval.

## Consent for publication

Not applicable.

## Authors’ contributions

M.A and Y.D wrote the code of the analysis and M.A. wrote the shiny app. M.A. and Y.D. wrote the main manuscript text, and prepared the figures and tables. G.K. and C.M. supervised the study. All authors reviewed the manuscript.

## Acknowledgements

Not applicable.

## References

[1] Ralph A DeFronzo, Ele Ferrannini, Leif Groop, Robert R Henry, William H Herman, Jens Juul Holst, Frank B Hu C Ronald Kahn, Itamar Raz, Gerald I Shulman, et al. Type 2 diabetes mellitus. Nature reviews Disease primers, 1(1):1–22, 2015.

[2] Etie Moghissi, Faramarz Ismail-Beigi, and Robin Cammarota Devine. Hypoglycemia: minimizing its impact in type 2 diabetes. Endocrine Practice, 19(3):526–535, 2013.

[3] Belinda P Childs, Nathaniel G Clark, Daniel J Cox, Philip E Cryer, et al. Defining and reporting hypoglycemia in diabetes: a report from the american diabetes association workgroup on hypoglycemia. Diabetes care, 28(5):1245, 2005.

[4] Philip E Cryer. Severe hypoglycemia predicts mortality in diabetes. Diabetes Care, 35(9):1814–1816, 2012.

[5] Rozalina G McCoy, Holly K Van Houten, Jeanette Y Ziegenfuss, Nilay D Shah, Robert A Wermers, and Steven A Smith. Increased mortality of patients with diabetes reporting severe hypoglycemia. Diabetes Care, 35(9):1897–1901, 2012.

[6] Erwin C Puente, Julie Silverstein, Adam J Bree, Daniel R Musikantow, David F Wozniak, Susan Maloney, Dorit Daphna-Iken, and Simon J Fisher. Recurrent moderate hypoglycemia ameliorates brain damage and cognitive dysfunction induced by severe hypoglycemia. Diabetes, 59(4):1055–1062, 2010.

[7] Candace M Reno, Dorit Daphna-Iken, Y Stefanie Chen, Jennifer VanderWeele, Krishan Jethi, and Simon J Fisher. Severe hypoglycemia–induced lethal cardiac arrhythmias are mediated by sympathoadrenal activation. Diabetes, 62(10):3570–3581, 2013.

[8] Quan Zou, Kaiyang Qu, Yamei Luo, Dehui Yin, Ying Ju, and Hua Tang. Predicting diabetes mellitus with machine learning techniques. Frontiers in Genetics, 9:515, 2018.

[9] Robert J Tanenberg, Christopher A Newton, and Almond J Drake III. Confirmation of hypoglycemia in the “dead-in-bed” syndrome, as captured by a retrospective continuous glucose monitoring system. Endocrine Practice, 16(2):244–248, 2010.

[10] Justin B Echouffo-Tcheugui, Arnaud D Kaze, Gregg C Fonarow, and Sam Dagogo-Jack. Severe hypoglycemia and incident heart failure among adults with type 2 diabetes. The Journal of Clinical Endocrinology & Metabolism, 107(3):e955–e962, 2022.

[11] T Skrivarhaug, H-J Bangstad, LC Stene, L Sandvik, KF Hanssen, and G Joner. Long-term mortality in a nationwide cohort of childhood-onset type 1 diabetic patients in norway. Diabetologia, 49:298–305, 2006.

[12] Benjamin Shickel, Patrick James Tighe, Azra Bihorac, and Parisa Rashidi. Deep ehr: a survey of recent advances in deep learning techniques for electronic health record (ehr) analysis. IEEE Journal of Biomedical and Health Informatics, 22(5):1589–1604, 2017.

[13] Guthrie S Birkhead, Michael Klompas, and Nirav R Shah. Uses of electronic health records for public health surveillance to advance public health. Annual review of public health, 36:345–359, 2015.

[14] Nonie Alexander, Daniel C Alexander, Frederik Barkhof, and Spiros Denaxas. Identifying and evaluating clinical subtypes of alzheimer’s disease in care electronic health records using unsupervised machine learning. BMC Medical Informatics and Decision Making, 21(1):1–13, 2021.

[15] Xiaoyi Raymond Gao, Marion Chiariglione, Ke Qin, Karen Nuytemans, Douglas W Scharre, Yi-Ju Li, and Eden R Martin. Explainable machine learning aggregates polygenic risk scores and electronic health records for alzheimer’s disease prediction. Scientific Reports, 13(1):450, 2023.

[16] Tianhao Li, Zhishun Wang, Wei Lu, Qian Zhang, and Dengfeng Li. Electronic health records based reinforcement learning for treatment optimizing. Information Systems, 104:101878, 2022.

[17] Jeffrey P Anderson, Jignesh R Parikh, Daniel K Shenfeld, Vladimir Ivanov, Casey Marks, Bruce W Church, Jason M Laramie, Jack Mardekian, Beth Anne Piper, Richard J Willke, et al. Reverse engineering and evaluation of prediction models for progression to type 2 diabetes: an application of machine learning using electronic health records. Journal of Diabetes Science and Technology, 10(1):6–18, 2016.

[18] Tao Zheng, Wei Xie, Liling Xu, Xiaoying He, Ya Zhang, Mingrong You, Gong Yang, and You Chen. A machine learning-based framework to identify type 2 diabetes through electronic health records. International Journal of Medical Informatics, 97:120–127, 2017.

[19] Binh P Nguyen, Hung N Pham, Hop Tran, Nhung Nghiem, Quang H Nguyen, Trang TT Do, Cao Truong Tran, and Colin R Simpson. Predicting the onset of type 2 diabetes using wide and deep learning with electronic health records. Computer Methods and Programs in Biomedicine, 182:105055, 2019.

[20] Sriram Ramgopal, Christopher M Horvat, Naveena Yanamala, and Elizabeth R Alpern. Machine learning to predict serious bacterial infections in young febrile infants. Pediatrics, 146(3), 2020.

[21] Dharanija Madhavan, Katarina Cuk, Barbara Burwinkel, and Rongxi Yang. Cancer diagnosis and prognosis decoded by blood-based circulating microrna signatures. Frontiers in genetics, 4:116, 2013.

[22] Jie Zhu, Jingxiang Wang, Xin Wang, Mingjing Gao, Bingbing Guo, Miaomiao Gao, Jiarui Liu, Yanqiu Yu, Liang Wang, Weikaixin Kong, et al. Prediction of drug efficacy from transcriptional profiles with deep learning. Nature Biotechnology, 39(11):1444–1452, 2021.

[23] Xue-bin Wang, Ning-hua Cui, and Xia’nan Liu. A novel 6-metabolite signature for prediction of clinical outcomes in type 2 diabetic patients undergoing percutaneous coronary intervention. Cardiovascular Diabetology, 21(1):1–15, 2022.

[24] Yixiang Deng, Lu Lu, Laura Aponte, Angeliki M Angelidi, Vera Novak, George Em Karniadakis, and Christos S Mantzoros. Deep transfer learning and data augmentation improve glucose levels prediction in type 2 diabetes patients. NPJ Digital Medicine, 4(1):109, 2021.

[25] Brandon Ballinger, Johnson Hsieh, Avesh Singh, Nimit Sohoni, Jack Wang, Geoffrey Tison, Gregory Marcus, Jose Sanchez, Carol Maguire, Jeffrey Olgin, et al. Deepheart: semi-supervised sequence learning for cardiovascular risk prediction. In Proceedings of the AAAI conference on artificial intelligence, volume 32, 2018.

[26] Jiang Wu, Yuan-Bo Diao, Meng-Long Li, Ya-Ping Fang, and Dai-Chuan Ma. A semisupervised learning based method: Laplacian support vector machine used in diabetes disease diagnosis. Interdisciplinary Sciences: Computational Life Sciences, 1:151–155, 2009.

[27] Juhyeon Kim and Hyunjung Shin. Breast cancer survivability prediction using labeled, unlabeled, and pseudo-labeled patient data. Journal of the American Medical Informatics Association, 20(4):613–618, 2013.

[28] Seung Mi Lee, Yonghyun Nam, Eun Saem Choi, Young Mi Jung, Vivek Sriram, Jacob S Leiby, Ja Nam Koo, Ig Hwan Oh, Byoung Jae Kim, Sun Min Kim, et al. Development of early prediction model for pregnancy-associated hypertension with graph-based semi-supervised learning. Scientific Reports, 12(1):15793, 2022.

[29] Shekoofeh Azizi, Laura Culp, Jan Freyberg, Basil Mustafa, Sebastien Baur, Simon Kornblith, Ting Chen, Nenad Tomasev, Jovana Mitrović, Patricia Strachan, et al. Robust and data-efficient generalization of self-supervised machine learning for diagnostic imaging. Nature Biomedical Engineering, pages 1–24, 2023.

[30] John B Buse, ACCORD Study Group, et al. Action to control cardiovascular risk in diabetes (accord) trial: design and methods. The American journal of Cardiology, 99(12):S21–S33, 2007.

[31] Tsang-Hsiang Cheng, Chih-Ping Wei, and Vincent S Tseng. Feature selection for medical data mining: Comparisons of expert judgment and automatic approaches. In 19th IEEE Symposium on Computer-Based Medical Systems (CBMS’06), pages 65–170. IEEE, 2006.

[32] Bandar Alotaibi and Munif Alotaibi. Consensus and majority vote feature selection methods and a detection technique for web phishing. Journal of Ambient Intelligence and Humanized Computing, 12:717–727, 2021.

[33] Max Kuhn. Building predictive models in r using the caret package. Journal of Statistical Software, 28:1–26, 2008.

[34] Leo Breiman. Random forests. Machine Learning, 45:5–32, 2001.

[35] Avrim Blum and Tom Mitchell. Combining labeled and unlabeled data with co-training. In Proceedings of the Eleventh Annual Conference on Computational Learning Theory, pages 92–100, 1998.

[36] Rie Kubota Ando and Tong Zhang. Two-view feature generation model for semi-supervised learning. In Proceedings of the 24th international conference on Machine learning, pages 25–32, 2007.

[37] Sham M Kakade and Dean P Foster. Multi-view regression via canonical correlation analysis. In Learning Theory: 20th Annual Conference on Learning Theory, COLT 2007, San Diego, CA, USA; June 13-15, 2007. Proceedings 20, pages 82–96. Springer, 2007.

[38] Shipeng Yu, Balaji Krishnapuram, Harald Steck, R Rao, and Rómer Rosales. Bayesian co-training. Advances in Neural Information Processing Systems, 20, 2007.

[39] Laurens Van der Maaten and Geoffrey Hinton. Visualizing data using t-sne. Journal of Machine Learning Research, 9(11), 2008.

[40] Robert Tibshirani. Regression shrinkage and selection via the lasso. Journal of the Royal Statistical Society: Series B (Methodological), 58(1):267–288, 1996.

[41] Miron B Kursa and Witold R Rudnicki. Feature selection with the boruta package. Journal of Statistical Software, 36:1–13, 2010.

[42] Hanchuan Peng, Fuhui Long, and Chris Ding. Feature selection based on mutual information criteria of max-dependency, max-relevance, and min-redundancy. IEEE Transactions on Pattern Analysis and Machine Intelligence, 27(8):1226–1238, 2005.

[43] Zhenyu Zhao, Radhika Anand, and Mallory Wang. Maximum relevance and minimum redundancy feature selection methods for a marketing machine learning platform. In 2019 IEEE International Conference on Data Science and Advanced Analytics (DSAA), pages 442–452. IEEE, 2019.

[44] Winston Chang, Joe Cheng, JJ Allaire, Carson Sievert, Barret Schloerke, Yihui Xie, Jeff Allen, Jonathan McPherson, Alan Dipert, and Barbara Borges. shiny: Web Application Framework for R, 2023. R package version 1.7.4.9002.

[45] Scott M Lundberg, Bala Nair, Monica S Vavilala, Mayumi Horibe, Michael J Eisses, Trevor Adams, David E Liston, Daniel King-Wai Low, Shu-Fang Newman, Jerry Kim, et al. Explainable machine-learning predictions for the prevention of hypoxaemia during surgery. Nature biomedical engineering, 2(10):749–760, 2018.

[46] N Hermanns, B Kulzer, T Kubiak, M Krichbaum, and T Haak. The effect of an education programme (hypos) to treat hypoglycaemia problems in patients with type 1 diabetes. Diabetes/metabolism Research and Reviews, 23(7):528–538, 2007.

[47] Eric L Johnson. Glycemic variability in type 2 diabetes mellitus: oxidative stress and macrovascular complications. Diabetes: An Old Disease, a New Insight, pages 139–154, 2013.

[48] Chen Long, Yaling Tang, Huang Jiang Sheng, Suo Liu, and Zhenhua Xing. Association of long-term visit-to-visit variability of hba1c and fasting glycemia with hypoglycemia in type 2 diabetes mellitus brief title: Variability of hba1c, fasting glycemia, and hypoglycemia. Frontiers in Endocrinology, page 1870, 2022.

[49] Diabetes Control and Complications Trial Research Group. The effect of intensive treatment of diabetes on the development and progression of long–term complications in insulin-dependent diabetes mellitus. New England journal of medicine, 329(14):977–986, 1993.

[50] Michael Fralick, David Dai, Chloe Pou-Prom, Amol A Verma, and Muhammad Mamdani. Using machine learning to predict severe hypoglycaemia in hospital. Diabetes, Obesity and Metabolism, 23(10):2311–2319, 2021.

