## Supplementary Material for "Long-term Prediction of Severe Hypoglycemia in Type 2 Diabetes Based on Multi-view Co-training"

### Supplementary Material (SM)

#### Candidate Risk Factors

Table 1: Comparison of the results obtained from a single-view co-training model using MD and MRMR feature selection data. Features of the MD data; FPG mean, FPG STD, HbA1c mean, HbA1c STD, g1check mean, g1check STD, g1diabed mean, g1diabed STD, g1nutrit mean, g1nutrit STD. Features of the MRMR data; FPG STD, HbA1c mean, g1diabed STD and NPHL insulin mean. Percentage=(X last iteration-X $1st$ iteration)×100/ (X $1st$ iteration). Based on the single-view co-training algorithm, we observe great improvement for the NB method in specificity, sensitivity, and accuracy.

| | NB | | | | | |
|---|---|---|---|---|---|---|
| | NPV | PPV | Spec | Sens | Acc | F1 |
| MD $1st$ iteration | 0.924 | 0.131 | 0.038 | 0.979 | 0.159 | 0.230 |
| MD last iteration | 0.900 | 0.127 | 0.133 | 0.865 | 0.228 | 0.221 |
| Percentage change | -2.60% | -2.63% | **249.21%** | -11.64 | **43.38%** | -3.86 |
| MRMR $1st$ iteration | 0.894 | 0.183 | 0.740 | 0.407 | 0.696 | 0.248 |
| MRMR last iteration | 0.868 | 0.129 | 0.265 | 0.740 | 0.326 | 0.219 |
| Percentage change | -2.89% | -29.94% | -64.17% | **81.76%** | -53.17% | -11.97% |

| | RF | | | | | |
|---|---|---|---|---|---|---|
| | NPV | PPV | Spec | Sens | Acc | F1 |
| MD $1st$ iteration | 0.920 | 0.185 | 0.570 | 0.663 | 0.582 | 0.289 |
| MD last iteration | 0.924 | 0.186 | 0.561 | 0.689 | 0.577 | 0.293 |
| Percentage change | (0.40%) | (0.85%) | -1.52% | **3.81%** | -0.87% | **1.39%** |
| MRMR $1st$ iteration | 0.916 | 0.176 | 0.548 | 0.658 | 0.562 | 0.277 |
| MRMR last iteration | 0.914 | 0.171 | 0.545 | 0.639 | 0.557 | 0.268 |
| Percentage change | -0.20% | -3.09% | -0.72% | -2.92% | -0.89% | -3.2% |

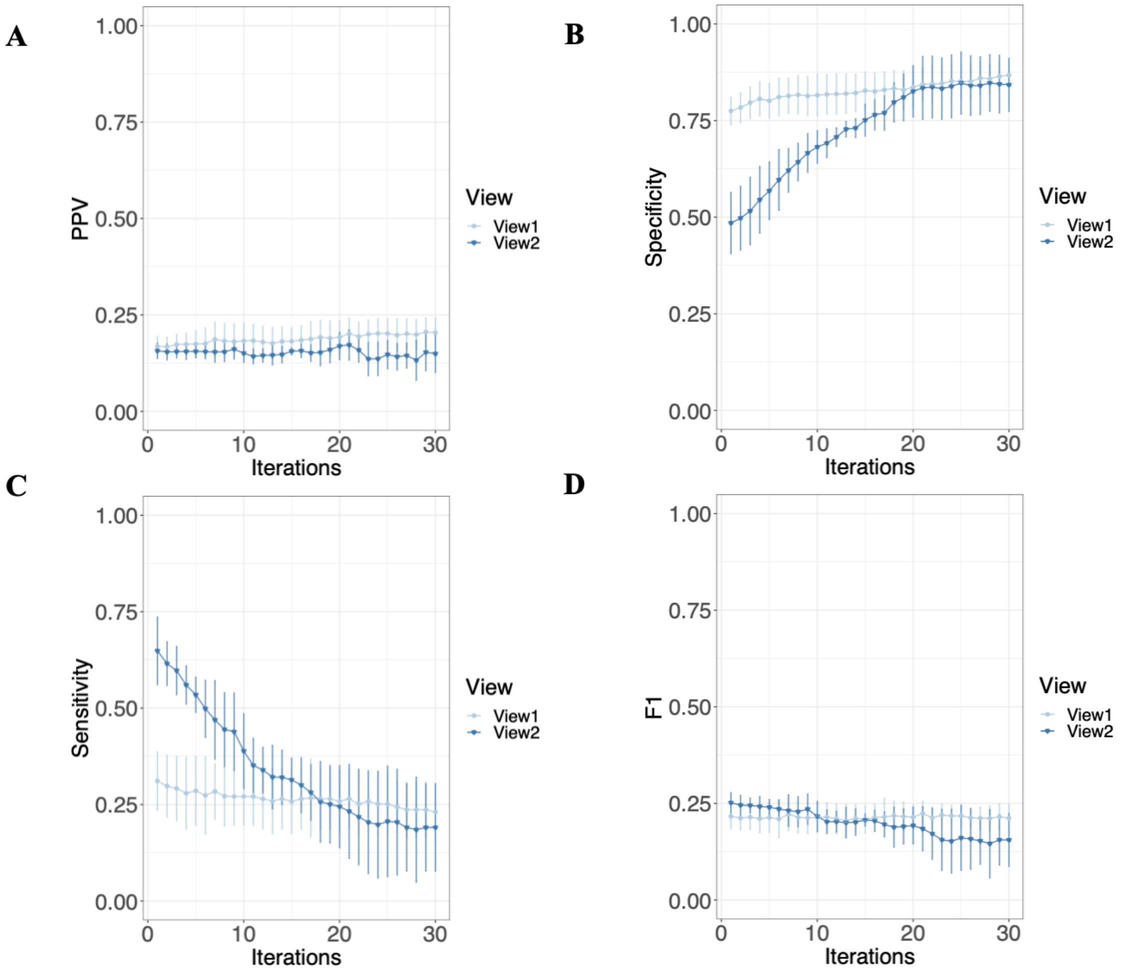


Figure 1: **Mean accuracy metrics of test results for each iteration in multi-view co-training for NB using MD features.** We split the data into two different labeled data sets View 1 and View 2. From the unlabeled data, we create two pools of datasets: U1 and U2. Following the training process, predictions are generated for both U1 and U2, and pseudo labels are created. The most confident labels are then added to the labeled dataset. The iteration is repeated thirty times. We observe a great improvement in specificity at the expense of accuracy and sensitivity. **(A)** Positive Predicted Value (PPV) or Precision **(B)** Specificity **(C)** Sensitivity **(D)** F1-score (F1).

Table 2: Results of combining views using both AND and OR rules. When we combine them in either way, RF wins, but it prevails in MD selection OR rule, while MRMR prevails in the AND rule. The AND rule yields higher PPV, specificity, and accuracy, whereas the OR rule yields higher NPV, sensitivity, and F1. More balanced specificity and sensitivity results are obtained with the OR rule.

AND Rule

| | NPV | PPV | Spec | Sens | Acc | F1 |
|---|---|---|---|---|---|---|
| NB-MD | 0.870 | 0.050 | 0.982 | 0.007 | 0.856 | 0.012 |
| NB-MRMR | 0.872 | 0.167 | 0.981 | 0.026 | 0.858 | 0.046 |
| RF-MD | 0.877 | 0.245 | 0.961 | 0.086 | 0.849 | 0.128 |
| RF-MRMR | 0.873 | **0.300** | **0.993** | 0.020 | **0.868** | 0.037 |

OR Rule

| | NPV | PPV | Spec | Sens | Acc | F1 |
|---|---|---|---|---|---|---|
| NB-MD | 0.892 | 0.179 | 0.728 | 0.404 | 0.686 | 0.248 |
| NB-MRMR | 0.889 | 0.183 | 0.770 | 0.351 | 0.716 | 0.241 |
| RF-MD | **0.906** | 0.173 | 0.584 | **0.589** | 0.585 | **0.267** |
| RF-MRMR | 0.885 | 0.161 | 0.715 | 0.371 | 0.671 | 0.224 |

Table 3: All candidate risk factors with folder names.

| | **Variables** | **Description** | **Type** |
|---|---|---|---|
| 1 | Female | Gender: 0=Male, 1=Female | Num |
| 2 | Baseline age | Age (yrs): Randomization | Num |
| 3 | Arm | Randomization Arm: 1-8 | Char |
| 4 | cvd_hx_baseline | CVD History at Baseline: 0=No, 1=Yes | Num |
| 5 | Raceclass | Race Class: White, Black, Hispanic, Other | Char |
| | | Data Set Name: accord_key.sas7bdat | |
| 6 | sbp | Systolic Blood Pressure (mmHg) | Num |
| 7 | dbp | Diastolic Blood Pressure (mmHg) every month in the first four months; after that every two months | Num |
| | | Data Set Name: bloodpressure.sas7bdat | |
| 8 | loop | Loop diuretics | Num |
| 9 | thiazide | Thiazide diuretics | Num |
| 10 | ksparing | Ksparing diuretics | Num |
| 11 | potassium | Potassium supplements | Num |
| 12 | a2rb | Angiotensin type 2 antagonists (ARB) | Num |
| 13 | acei | Ace inhibitors | Num |

Continued on next page

Table 3 – continued from previous page

| | Variables | Description | Type |
|---|---|---|---|
| 14 | dhp_ccb | Dihydropyridine calcium channel blockers | Num |
| 15 | nondhp_ccb | Non-dihydropyridine calcium channel blockers | Num |
| 16 | alpha_blocker | Peripheral alpha-blockers | Num |
| 17 | central_agent | Central alpha-adrenergic agonists | Num |
| 18 | beta_blocker | Beta-blockers | Num |
| 19 | vasodilator | Vasodilators | Num |
| 20 | reserpine | Reserpine | Num |
| 21 | other_bpmed | Other antihypertensive agents | Num |
| 22 | digitalis | Digitalis preparations | Num |
| 23 | antiarrhythmic | Anti-arrhythmics | Num |
| 24 | nitrate | Nitrates | Num |
| 25 | other_cvmed | Other cardiovascular drugs | Num |
| 26 | sulfonylurea | Sulfonylureas | Num |
| 27 | biguanide | Biguanides | Num |
| 28 | meglitinide | Meglitinides | Num |
| 29 | ag_inhibitor | Alpha-glucosidase inhibitors | Num |
| 30 | nphl_insulin | NPH or L Insulins | Num |
| 31 | tzd | Thiazolidinediones | Num |
| 32 | reg_insulin | Regular Insulins | Num |
| 33 | la_insulin | Lispro or Aspart Insulins | Num |
| 34 | othbol_insulin | Other Bolus Insulins | Num |
| 35 | premix_insulin | Premixed Insulins | Num |
| 36 | other_diabmed | Other Diabetes Treatments | Num |
| 37 | bile_sequestrant | Bile-acid sequestrants | Num |
| 38 | statin | HMG CoA reducatce inhibitors (statins) | Num |
| 39 | fibrate | Fibrates | Num |
| 40 | other_lipidmed | Miscellaneous lipid-lowering drugs | Num |
| 41 | cholest_abi | Cholesterol absorption inhibitors | Num |
| 42 | niacin | Niacin and nicotinic acid | Num |
| 43 | anti_coag | Oral anticoagulants (warfarin, coumadin, anisindione) | Num |
| 44 | anti_inflam | Non-steroidal anti-inflammatory agents (excluding aspirin) | Num |
| 45 | platelet_agi | Inhibitors of platelet aggregation (except aspirin) | Num |
| 46 | cox2 | Cox-2 inhibitors | Num |
| 47 | aspirin | Aspirin | Num |
| 48 | thyroid | Thyroid agents | Num |
| 49 | progestin | Progestins | Num |
| 50 | estrogen | Estrogens (excluding vaginal creams) | Num |
| 51 | oral_asthma | Oral asthma drugs (except steroids) | Num |
| 52 | anti_depress | Any antidepressant | Num |

Continued on next page

Table 3 – continued from previous page

| | Variables | Description | Type |
|---|---|---|---|
| 53 | inhaled_asthma | Inhaled steroids for asthma | Num |
| 54 | oral_steroid | Oral steroids | Num |
| 55 | anti_psych | Any antipsychotic | Num |
| 56 | osteoporosis | Drugs for osteoporosis | Num |
| 57 | fluid_retention | Diuretic for fluid retention | Num |
| 58 | other_med | Any other prescribed medication | Num |
| 59 | vitamin | Vitamins and/or nutritional supplements | Num |
| 60 | otc | Over-the-counter medications | Num |
| | | Data Set Name: concomitantmeds.sas7bdat | |
| 61 | Hba1c | glycosylated hemoglobin (%) | Num |
| | | Data Set Name: hba1.sas7bdat | |
| 62 | chol | Total Cholesterol (mg/dL) | Num |
| 63 | trig | Triglycerides (mg/dL) | Num |
| 64 | hdl | High density lipoprotein (mg/dL) | Num |
| | | Data Set Name: lipids.sas7bdat | |
| 65 | Total_Brain _Volume_ICV | Intracranial Volume (cc, algorithm) | Num |
| 66 | Gray_Matter _total_sum | Gray Matter, Total (cc, summed) | Num |
| 67 | white_matter _total_sum | White Matter, Total (cc, summed) | Num |
| | | Data Set Name: mind_mri.sas7bdat | |
| 68 | FPG | Fasting plasma glucose (mg/dL) | Num |
| 69 | Serum creatinine | Serum creatinine (mg/dL) | Num |
| 70 | eGFR | eGFR from 4 variable MDRD equation (ml/min/1.73 $m^2$) | Num |
| 71 | UALB | Urinary albumin (mg/dL) | Num |
| 72 | UACR | Urinary albumin to creatinine ratio (mg/g) | Num |
| | | Data Set Name: otherlabs.sas7bdat | |
| 73 | livealon | participant lives with one or more adults | Num |
| 74 | edu | participant's highest level of education | Num |
| 75 | yrsdiab | year of diabetes diagnosis | Num |
| 76 | yrslipi | year of hyperlipidemia diagnosis | Num |
| 77 | yrstens | year of hypertension diagnosis | Num |
| 78 | ulcer | foot ulcer requiring antibiotics | Num |
| 79 | protein | protein in urine | Num |
| 80 | hartfail | heart failure/CHF | Num |
| 81 | neuropat | neuropathy/nerve problems | Num |
| 82 | depressn | depression | Num |
| 83 | histhart | family history of heart disease, heart attack, or stroke | Num |

Continued on next page

Table 3 – continued from previous page

| | Variables | Description | Type |
|---|---|---|---|
| 84 | cigarett | smoked cigarettes in last 30 days | Num |
| 85 | smokelif | smoked more than 100 cigarettes during lifetime | Num |
| 86 | alcohol | number of alcoholic drinks consumed weekly | Num |
| 87 | waist_cm | waist circumference (cm) | Num |
| 88 | scrright | visual acuity score, right eye | Num |
| 89 | scrleft | visual acuity score, left eye | Num |
| 90 | retpathy | participant had retinopathy | Num |
| 91 | visloss | vision loss | Num |
| 92 | fankle | ankle reflexes | Num |
| 93 | BMI | Body mass index (kg/m$^2$) | Num |
| | | Data Set Name: f07_baselinehistoryphysicalexam.sas7bdat | |
| 94 | g1nutrit | nutrition education, time in minutes | Num |
| 95 | g1diabed | general diabetes education (other than nutrition), time in minutes | Num |
| 96 | g1check | average frequency of blood sugar check, times per week, since last call or visit | Num |
| 97 | g17days | number of hypoglycemic episodes (SMBG ¡70 mg/dL or ¡3.9 mmol/L) in last 7 days | Num |
| 98 | g1_report | hypoglycemia requiring hospitalization, emergency care without hospital admission, or medical assistance without emergency care or hospitalization, number of times since last call or visit | Num |
| 99 | g2hwofch | frequency of use of CHO/Insulin ratio | Num |
| 100 | g2hwofba | frequency of insulin injection/inhalation as prescribed for basal (background) insulin | Num |
| 101 | g2hwofbo | frequency of insulin injection/inhalation as prescribed for injected bolus (premeal) insulin | Num |
| 102 | g2aveba | visit entry total basal insulin/day | Num |
| 103 | g2avebol | visit entry total bolus insulin/day | Num |
| 104 | g2avetid | visit entry total injected insulins/day | Num |
| | | Data Set Name: f08_09_glycemiamanagement.sas7bdat | |
| 105 | HoursAll | average hrs/week spent in any activity | Num |

Continued on next page

Table 3 – continued from previous page

| | Variables | Description | Type |
|---|---|---|---|
| 106 | HoursMod | average hrs/week spent in moderate activity | Num |
| | | Data Set Name: f29_champsphysicalactivity.sas7bdat | |
| 107 | x3malb | micro or macro albuminuria within past 2 years | Num |
| | | Data Set Name: f01_inclusionexclusionsummary.sas7bdat | |
| 108 | total_mmse | MMSE Total Score | Num |
| 109 | total_dsc | DSST Total Score | Num |
| 110 | stroop | STROOP Interference Score | Num |
| 111 | ravlt | RAVLT Score | Num |
| | | Data Set Name: mind.sas7bdat | |
| 112 | ses03 | participant experienced being thirsty during the past month | Num |
| 113 | Ses51 | participant experienced pains in legs or calves when walking during the past month | Num |
| | | Data Set Name: f23_hrql.sas7bdat | |
| 114 | ateskfat | how often participant ate raw vegetables or fresh fruit when snacking | Num |
| 115 | Aterdmt | participant ate red meat such as beef, pork, or lamb in last 3 months | Num |
| 116 | Substitution | DIET Factor 1: Average score for low-fat choices | Num |
| | | Data Set Name: f26_dietquestionnaire.sas7bdat | |
| 117 | HUI3Scor | Health Utilities Index Mark3 (HUI3); Aggregate score of vision, hearing, speech, ambulation, dexterity, emotion, cognition and pain | Num |
| 118 | HUI2pf | Health Utilities Index Mark2 (HUI2); Aggregate score of sensation, mobility, cognition, self-care, emotion, pain and fertility | Num |
| | | Data Set Name: f19_healthutilitiesindex.sas7bdat | |

Table 4: Characteristics of selected features according to follow-up in ACCORD dataset. mean ± standard deviation for continuous and count (percentage %) for the categorical variables.

| Demographics | Values |
|---|---|
| Age | $62.70 \pm 6.59$ |
| BMI ($kg/m^2$) | $32.22 \pm 5.39$ |
| Female | 496 (42.8%) |
| Fasting Glucose | $175.28 \pm 56.11$ |
| HbA1c | $8.30 \pm 1.05$ |
| g1check | $18.57 \pm 6.46$ |
| g1diabed | $18.46 \pm 12.30$ |
| g1nutrit | $13.09 \pm 9.63$ |
| Sulfonylurea | 520 (44.22%) |
| Meglitinide | 31 (2.64%) |
| Reg Insulin | 239 (20.32%) |
| La Insulin | 178 (15.14%) |
| Othbol Insulin | 44 (3.74%) |
| Premix Insulin | 212 (18.03%) |
| NPH or L Insulin | 594 (50.51%) |

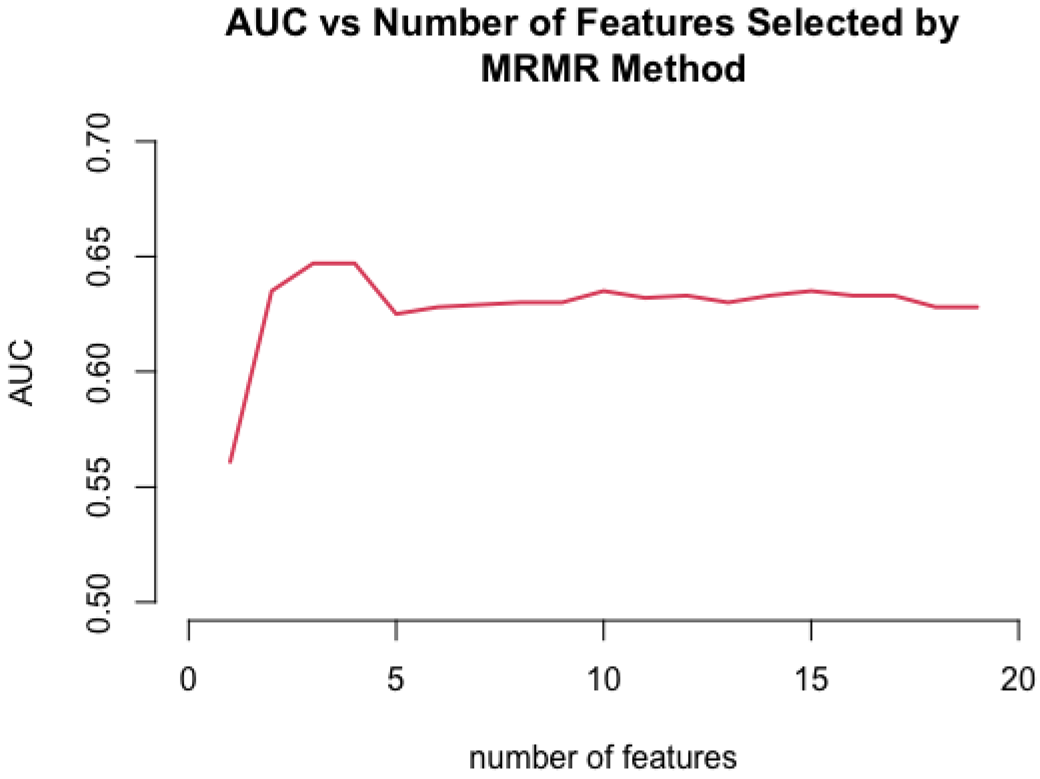


Figure 2: Area Under Curve (AUC) versus selected features by MRMR method. The number of features should be specified before running the MRMR code for entire ACCORD data. Therefore, we need to define how many features should be selected by MRMR method in advance. To do this, MRMR selected features from 1 to 17, then we calculated the AUC value for each. Finally, we obtained the highest AUC for the number of 4 features.

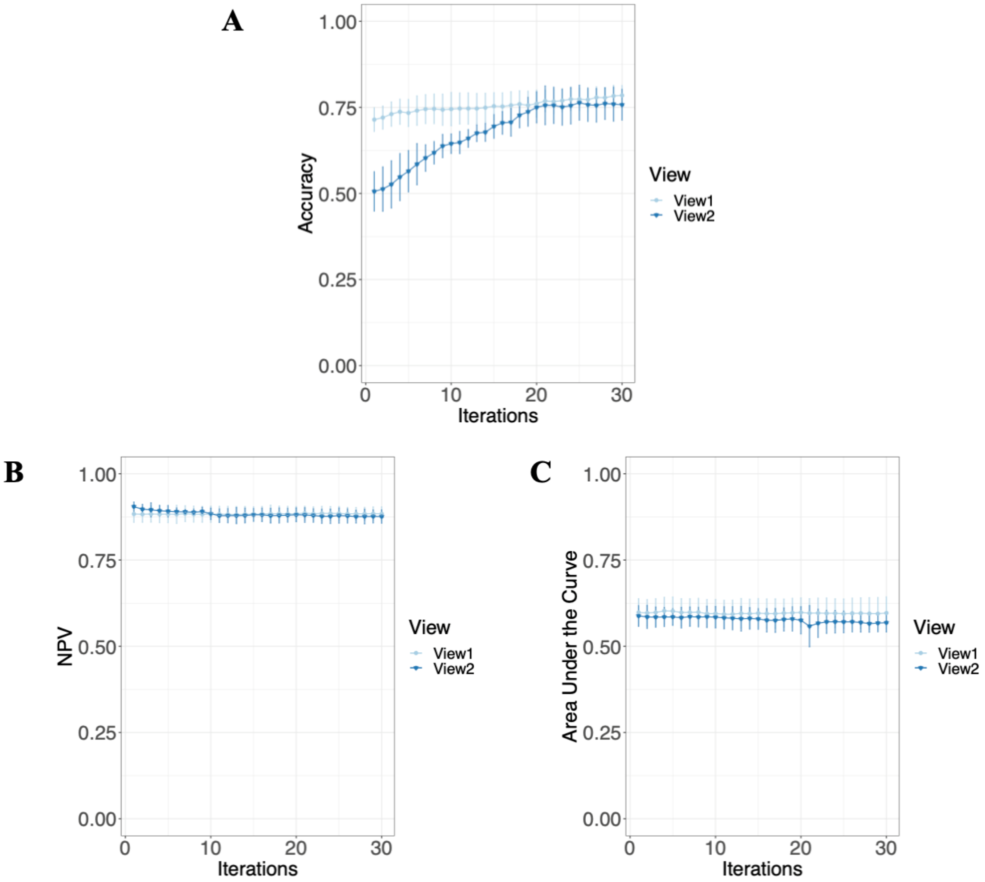


Figure 3: Mean accuracy metrics of test results for each iteration in multi-view co-training for NB MD features. We split the data into two different labeled data as View 1 and View 2. From the unlabeled data, we created two pools of datasets: U1 and U2. Following the training process, predictions are generated for both U1 and U2, and pseudo labels are created. The iteration repeated thirty times. (5 negative and 1 positive) (A) Accuracy, (B) NPV, (C) AUC-ROC

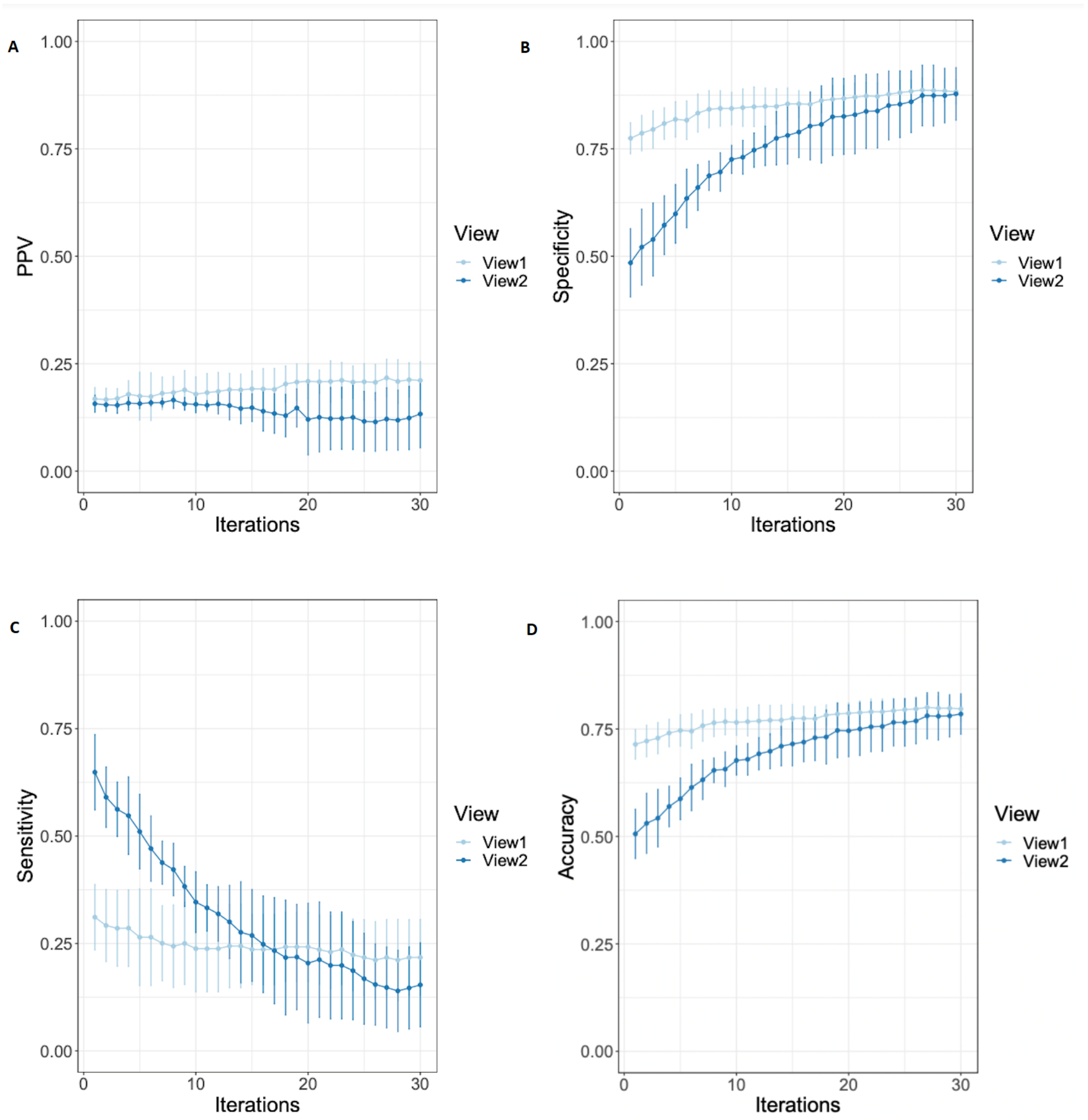


Figure 4: Mean accuracy metrics of test results for each iteration in multi-view co-training for NB MD features. We split the data into two different labeled data as View 1 and View 2. From the unlabeled data, we created two pools of datasets: U1 and U2. Following the training process, predictions are generated for both U1 and U2, and pseudo labels are created. The iteration repeated thirty times. (7 negative and 1 positive) (a) PPV, (b) Specificity, (c) Sensitivity, (d) Accuracy.

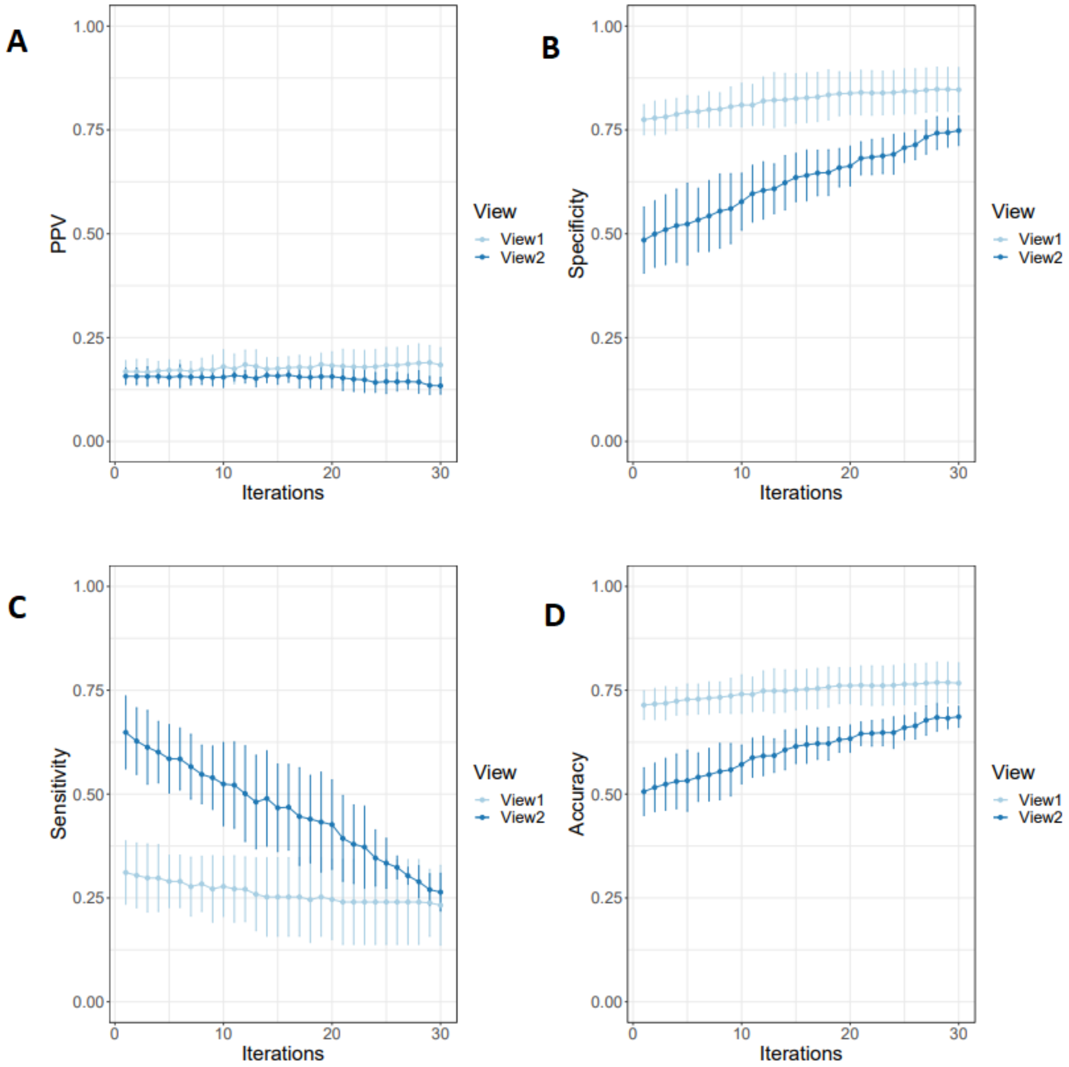


Figure 5: Mean accuracy metrics of test results for each iteration in multi-view co-training for NB MD features. We split the data into two different labeled data as View 1 and View 2. From the unlabeled data, we created two pools of datasets: U1 and U2. Following the training process, predictions are generated for both U1 and U2, and pseudo labels are created. The iteration repeated thirty times. (3 negative and 1 positive) (a) PPV, (b) Specificity, (c) Sensitivity, (d) Accuracy.

Table 5: Comparing multi-view co-training model results on MD and MRMR features with undersampling imbalanced solution MD Variables (View 1: Fpg Mean, Fpg Std, Hba1c Mean, Hba1c Std, View 2: g1check Mean, g1check Std, g1diabed Mean, g1diabed Std, g1nutrit Mean, g1nutrit Std), and MRMR (View 1: Fpg Std, Hba1c Mean, View 2: g1diabed Std, nphl insulin Mean) (7 negative and 1 positive).

| | NB | | | | | | | | | | | |
|---|---|---|---|---|---|---|---|---|---|---|---|---|
| | MD | | | | | | MRMR | | | | | |
| Iterations | NPV | PPV | Spec | Sens | Acc | F | NPV | PPV | Spec | Sens | Acc | F |
| View1 1st iteration | 0.883 | 0.168 | 0.775 | 0.311 | 0.714 | 0.217 | 0.887 | 0.192 | 0.804 | 0.314 | 0.740 | 0.233 |
| View1 last iteration | 0.884 | 0.211 | 0.883 | 0.217 | 0.797 | 0.211 | 0.883 | 0.202 | 0.880 | 0.212 | 0.793 | 0.204 |
| Percentage | (0.07%) | (25.45%) | (13.96%) | (-30.11%) | (-11.54%) | (2.71%) | (-0.52%) | (5.23%) | (9.43%) | (-32.36%) | (7.24%) | (-12.64%) |
| View2 1st iteration | 0.905 | 0.157 | 0.485 | 0.649 | 0.506 | 0.252 | 0.910 | 0.172 | 0.570 | 0.609 | 0.575 | 0.267 |
| View3 last iteration | 0.876 | 0.133 | 0.878 | 0.154 | 0.785 | 0.177 | 0.876 | 0.146 | 0.887 | 0.141 | 0.793 | 0.174 |
| Percentage | (-3.23%) | (-15.17%) | (81.07%) | (-76.31%) | (55.15%) | (-29.81%) | (-3.74%) | (-15.17%) | (55.65%) | (-76.89%) | (-37.88%) | (-34.98%) |

| | MD | | | | | | MRMR | | | | | |
|---|---|---|---|---|---|---|---|---|---|---|---|---|
| Iterations | NPV | PPV | Spec | Sens | Acc | F | NPV | PPV | Spec | Sens | Acc | F |
| View1 1st iteration | 0.898 | 0.162 | 0.565 | 0.568 | 0.565 | 0.251 | 0.899 | 0.159 | 0.544 | 0.586 | 0.549 | 0.249 |
| View1 last iteration | 0.887 | 0.209 | 0.843 | 0.276 | 0.770 | 0.237 | 0.886 | 0.187 | 0.812 | 0.295 | 0.745 | 0.227 |
| Percentage | (-1.27%) | (28.60%) | (49.04%) | (-51.39%) | (36.29%) | (-5.53%) | (-1.37%) | (17.65%) | (49.15%) | (-49.61%) | (35.59%) | (-8.94%) |
| View2 1st iteration | 0.905 | 0.162 | 0.528 | 0.618 | 0.540 | 0.256 | 0.931 | 0.161 | 0.393 | 0.785 | 0.444 | 0.266 |
| View2 last iteration | 0.879 | 0.183 | 0.879 | 0.183 | 0.789 | 0.180 | 0.872 | NA | 1.000 | 0.000 | 0.872 | NA |
| Percentage | (-2.85%) | (13.18%) | (66.64%) | (-70.46%) | (46.14%) | (-29.84%) | (-6.42%) | NA | (154.60%) | (-100.00%) | (96.36%) | NA |

Table 6: Results of combining views using both AND, and OR rules selected for 7 Negative and 1 positive pseudo labels.

| | AND Rule | | | | | |
|---|---|---|---|---|---|---|
| | NPV | PPV | Spec | Sens | Acc | F |
| NB MD | 0.872 | 0.143 | 0.988 | 0.013 | 0.863 | 0.024 |
| NB MRMR | 0.871 | 0.091 | 0.981 | 0.013 | 0.856 | 0.023 |
| RF MD | 0.874 | 0.240 | 0.982 | 0.040 | 0.861 | 0.068 |
| RF MRMR | 0.872 | NA | 1.000 | 0.000 | 0.872 | NA |

| | OR Rule | | | | | |
|---|---|---|---|---|---|---|
| | NPV | PPV | Spec | Sens | Acc | F |
| NB MD | 0.890 | 0.185 | 0.773 | 0.351 | 0.719 | 0.243 |
| NB MRMR | 0.890 | 0.190 | 0.787 | 0.338 | 0.730 | 0.243 |
| RF MD | 0.896 | 0.193 | 0.742 | 0.417 | 0.701 | 0.264 |
| RF MRMR | 0.886 | 0.186 | 0.812 | 0.291 | 0.745 | 0.227 |

Table 7: Comparing multi-view co-training model results on MD and MRMR selected data with undersampling imbalanced solution. Results of the MD data (View 1: Fpg Mean, Fpg Std, Hba1c Mean, Hba1c Std, View 2: g1check Mean, g1check Std, g1diabed Mean, g1diabed Std, g1nutrit Mean, g1nutrit Std) and MRMR data (View 1: Fpg Std, Hba1c Mean, View 2: g1diabed Std, nphl insulin Mean) (3 negative and 1 positive ).

| | NB | | | | | | | | | | | |
|---|---|---|---|---|---|---|---|---|---|---|---|---|
| | MD | | | | | | MRMR | | | | | |
| Iterations | NPV | PPV | Spec | Sens | Acc | F | NPV | PPV | Spec | Sens | Acc | F |
| View1 1st iteration | 0.883 | 0.168 | 0.775 | 0.311 | 0.714 | 0.217 | 0.887 | 0.192 | 0.804 | 0.314 | 0.740 | 0.233 |
| View1 last iteration | 0.881 | 0.184 | 0.847 | 0.232 | 0.767 | 0.200 | 0.883 | 0.192 | 0.845 | 0.247 | 0.767 | 0.210 |
| Percentage | (-0.24%) | (9.35%) | (9.302%) | (-25.30%) | (7.38%) | (-7.41%) | (-0.516%) | (-0.06%) | (5.06%) | (-21.35%) | (3.68%) | (-9.92%) |
| View2 1st iteration | 0.905 | 0.157 | 0.485 | 0.649 | 0.506 | 0.252 | 0.910 | 0.172 | 0.570 | 0.609 | 0.575 | 0.267 |
| View2 last iteration | 0.874 | 0.134 | 0.748 | 0.264 | 0.686 | 0.177 | 0.883 | 0.141 | 0.722 | 0.340 | 0.673 | 0.198 |
| Percentage | (-3.45%) | (-14.62%) | (54.36%) | (-59.31%) | (35.65%) | (-29.80%) | (-2.96%) | (-18.06%) | (26.60%) | (-44.14%) | (17.16%) | (-26.02%) |
| | MD | | | | | | MRMR | | | | | |
| Iterations | NPV | PPV | Spec | Sens | Acc | F | NPV | PPV | Spec | Sens | Acc | F |
| 1st iteration | 0.898 | 0.162 | 0.564 | 0.568 | 0.564 | 0.251 | 0.897 | 0.157 | 0.543 | 0.579 | 0.548 | 0.246 |
| Last | 0.894 | 0.177 | 0.698 | 0.444 | 0.664 | 0.252 | 0.883 | 0.148 | 0.659 | 0.407 | 0.626 | 0.216 |
| Percentage | (-0.46%) | (9.04%) | (23.60%) | (-21.83%) | (17.79%) | (0.45%) | (-1.61%) | (-5.53%) | (21.19%) | (-29.70%) | (14.29%) | (-12.13%) |
| View1 1st iteration | 0.905 | 0.162 | 0.529 | 0.618 | 0.541 | 0.256 | 0.931 | 0.161 | 0.392 | 0.785 | 0.443 | 0.266 |
| View1 last iteration | 0.887 | 0.162 | 0.678 | 0.418 | 0.645 | 0.233 | 0.879 | 0.123 | 0.785 | 0.266 | 0.717 | 0.209 |
| Percentage | (-1.95%) | (0.185%) | (28.32%) | (-32.35%) | (19.19%) | (-9.243%) | (-5.62%) | (-23.75%) | (100.27%) | (-66.10%) | (61.79%) | (-21.55%) |

Table 8: Results of combining views using both AND, and OR rules selected for 3 Negative and 1 positive pseudo labels.

| | AND Rule | | | | | |
|---|---|---|---|---|---|---|
| | NPV | PPV | Spec | Sens | Acc | F |
| NB MD | 0.872 | 0.128 | 0.960 | 0.040 | 0.842 | 0.061 |
| NB MRMR | 0.876 | 0.194 | 0.943 | 0.093 | 0.834 | 0.126 |
| RF MD | 0.882 | 0.209 | 0.900 | 0.179 | 0.808 | 0.193 |
| RF MRMR | 0.880 | 0.214 | 0.921 | 0.146 | 0.821 | 0.173 |
| | OR Rule | | | | | |
| | NPV | PPV | Spec | Sens | Acc | F |
| NB MD | 0.887 | 0.154 | 0.635 | 0.450 | 0.611 | 0.229 |
| NB MRMR | 0.892 | 0.161 | 0.623 | 0.490 | 0.606 | 0.242 |
| RF MD | 0.910 | 0.161 | 0.475 | 0.682 | 0.502 | 0.260 |
| RF MRMR | 0.880 | 0.137 | 0.522 | 0.517 | 0.521 | 0.217 |

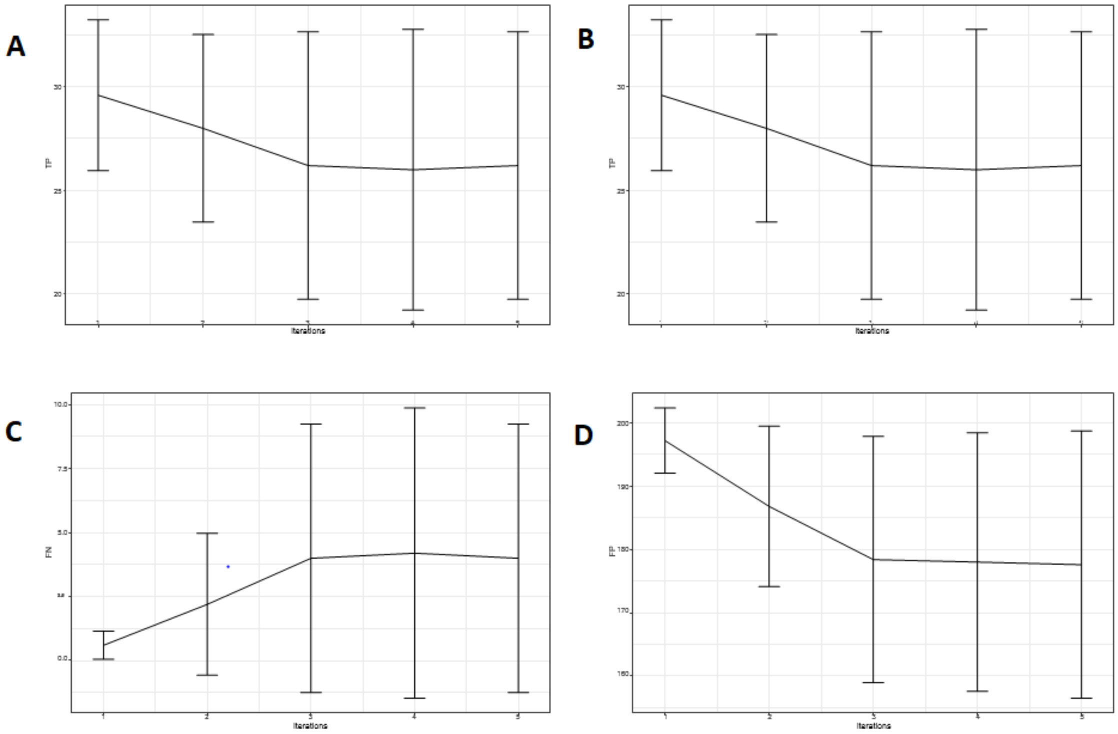


Figure 6: Confusion matrix for single-view co-training (A) TP, (B) TN, (C) FN, (D)FP.

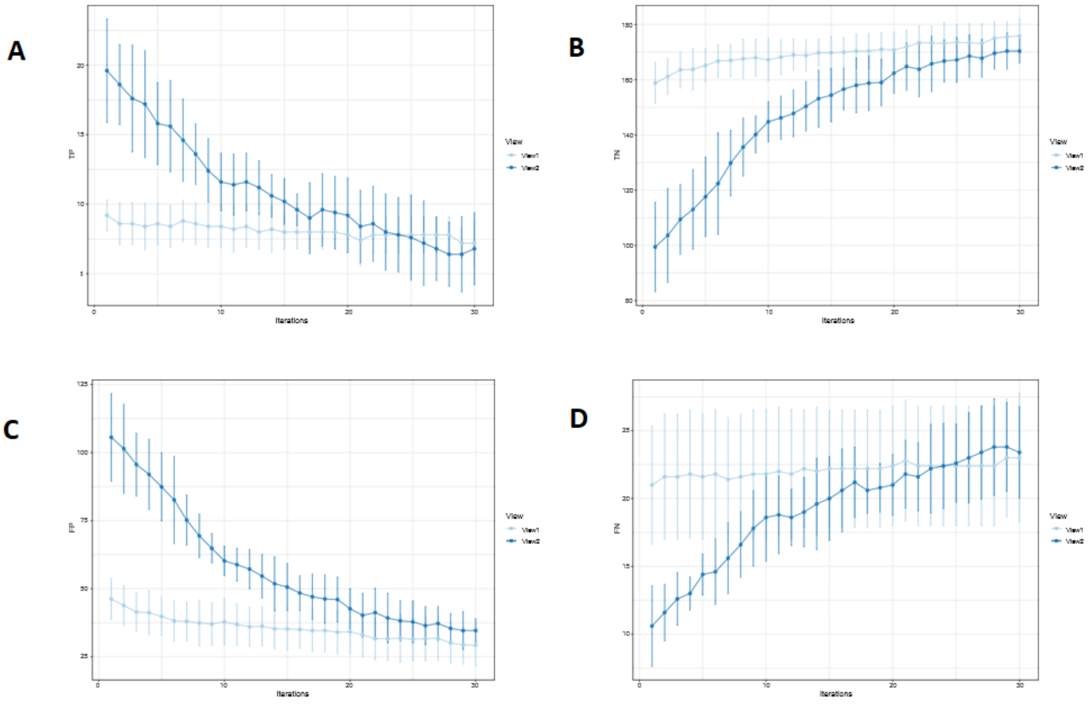


Figure 7: Confusion matrix for multi-view co-training (A) TP, (B) TN, (C) FN, (D) FP.

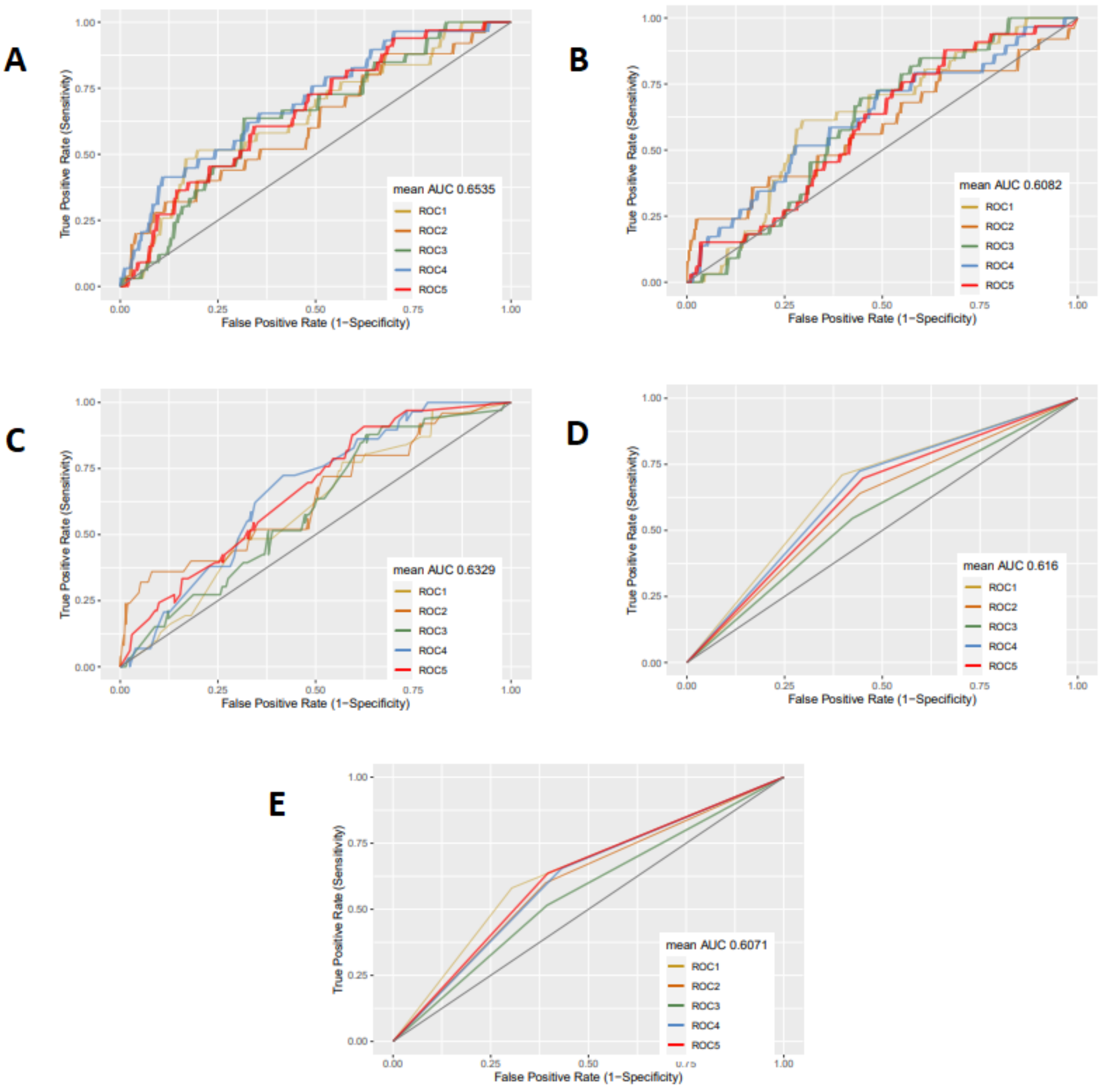


Figure 8: 5-fold ROC curves of conventional machine learning algorithms for (A) LR, (B) NB, (C) XGBoost, (D) RF, (E) SVM.

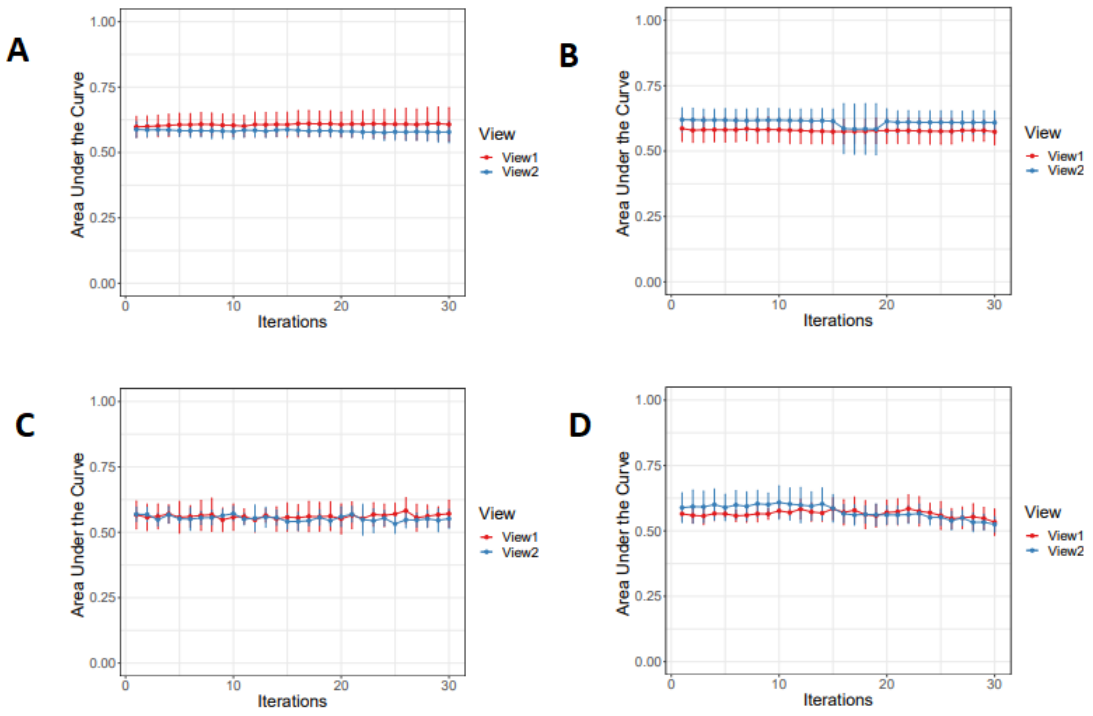


Figure 9: 5 fold AUC-ROC curves of multi-view co-training algorithm (5 negative and 1 positive) (a):NB MD (View 1 first AUC result: 0.5980, View 1 30th AUC result: 0.6075; View 2 first AUC result: 0.5879 and View 2 30th AUC result: 0.5784) , (b):NB MRMR(View 1 first AUC result: 0.5980 and View 1 30th AUC result: 0.6075; View 2 first AUC result: 0.5879 and View 2 30th AUC result: 0.5784) (c): RF MD (View 1 first AUC result: 0.5665 and View 1 30th AUC result: 0.5711; View 2 first AUC result: 0.5879 and View 2 30th AUC result: 0.5784) (d): RF MRMR(View 1 first AUC result: 0.5653 and View 1 30th AUC result: 0.5334; View 2 first AUC result: 0.5889 and View 2 30th AUC result: 0.5249).

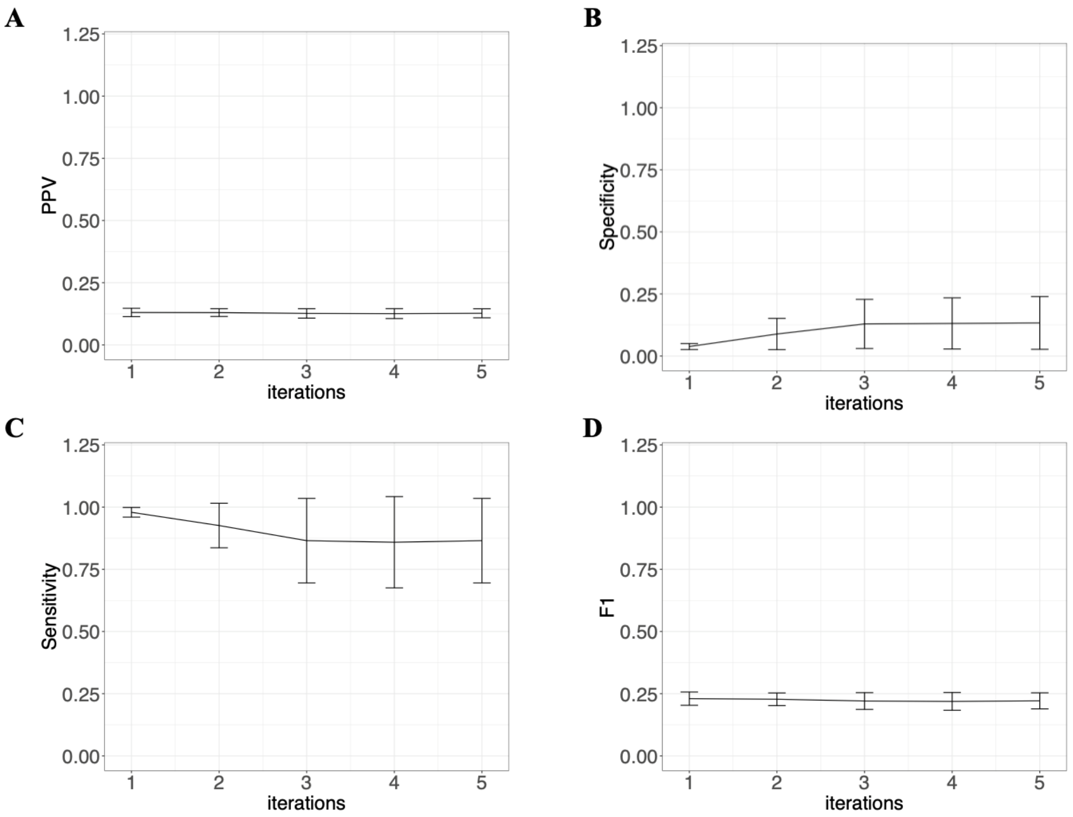


Figure 10: Mean accuracy metrics of test results for each iteration in single-view co-training for NB using MD features. The error bars show the mean ± standard deviations. We first split the labeled data as train and test set, then train the labeled data with the naive Bayes model and after that, we create the pseudo labels by estimating the unlabeled data and selecting the most confident labels, corresponding to probabilities higher than 0.9. This procedure is repeated until there is no unlabeled data to be selected. The figure shows the performance metrics of the test data results as (A) Positive Predicted Value (PPV), (B) Specificity, (C) Sensitivity, (D) F1-score (F1).

Table 9: Main features: The 17 features used in our study.

| Variables | Description |
|---|---|
| hba1c mean | Mean of HbA1c values |
| hba1c std | Standard deviation of HbA1c values |
| fpg mean | Mean of fasting plasma glucose values |
| fpg std | Standard deviation of fasting plasma glucose values |
| g1check mean | Mean of average frequency of blood sugar check |
| g1check std | Standard deviation of average frequency of blood sugar check |
| g1diabed mean | Mean of general diabetes education |
| g1diabed std | Standard deviation of general diabetes education |
| g1nutrit mean | Mean of nutrition education |
| g1nutrit std | Standard deviation of nutrition education |
| sulfonylurea mean | Mean of sulfonylureas |
| meglitinide mean | Mean of meglitinides |
| nphl mean | Mean of NPH or L insulins |
| reg insulin mean | Mean of regular insulins |
| la insulin mean | Mean of lispro or aspart insulins |
| othbol insulin mean | Mean of other bolus insulins |
| premix insulin mean | Mean of premixed insulins |

#### Feature Selection Methods

We first created all 118 candidate risk factors of SH events, which are listed in the supplemental Table 9. Afterwards, a down selection of the most significant variables was made by medical doctors from the Endocrinology Department at Beth Israel Medical Center. The doctors selected the top-12 features out of the 118 candidate risk factors. As the ACCORD study is a follow-up study, we computed the mean and standard deviations of the observations. This process resulted in 17 variables for further analysis, as represented in Table 9.

##### Least Absolute Shrinkage and selection operator (LASSO)

LASSO is a regression technique that uses the following loss function for both regularization and feature selection,

$$\text{Loss} = \sum_{i=1}^{N} \left( \left| y_i - \sum_j \beta_j x_{ij} \right|^2 + \lambda \sum_j \beta_j \right) \tag{1}$$

where $\lambda \sum_j \beta_j < t$ is the $L_1$ norm of the $\beta$ parameter vector and $\lambda$ is a non-negative parameter $\sum_j \beta_j$ controlling the degree of regularization. The ACCORD dataset consists of 118 variables. We created a potential estimation variable list by selecting 12 variables from the ACCORD dataset. Subsequently, we applied the LASSO feature selection to the dataset and compared the results obtained from these variables with those from the MD data and the other feature selection algorithm results. Finally, the features selected according to this method are FPG std, HbA1c mean, g1diabed std, NPHL Insulin, FPG mean, g1nutrit std, Othbol Insulin, Sulfonylurea mean, and Premix Insulin mean.

##### Boruta

Boruta is a feature selection method that uses a wrapper method built in the RF classifier. The Boruta method selects the features by following the steps below. 1.Synthetic new variables, called shadow features, are generated by randomly shuffling each variable. Shadow features are concatenated with the original dataset. 2.RF algorithm is run on X on y and Z scores are computed. 3.Calculate the maximum Z-score (MZSA) within the shadow features. 4.Original variables that are significantly lower than this MZSA are marked as unimportant. Original variables that are significantly higher than MZSA are marked as important. 5.Shadow features are deleted. 6. The algorithm is repeated until all variables are marked as important or unimportant, or until the constraint of the RF algorithm persists. We applied the Boruta feature selection method to the data and compared the results obtained from these variables with those from the MD data. Finally, the features selected according to this method are FPG std, HbA1c mean, g1diabed std, NPHL Insulin, FPG mean, g1nutrit std, Othbol Insulin, and g1 nutrition mean.

#### Maximum Relevance-Minimum Redundancy (MRMR).

This method maximizes the relevance of selected features while minimizing their redundancy. Since FCQ method in MRMR is the simplest, fastest and more effective than other methods, we focus on this method in this article. The MRMR method first calculates the maximum relevance between X and y and minimum redundancy between covariates itself, then by dividing the relevance to redundancy it calculates the FCQ score. Relevance is computed by the $F(X, y)$-statistics between X and y. Redundancy is calculated by correlation between the $X_i$. So FCQ score ($f^{FCQ}$) can be calculated by the following formula:

$$f^{FCQ} = \frac{F(X_i, y)}{\frac{1}{|S|}\sum_{X_s \in S} \rho(X_s, X_i)} \quad (2)$$

1. Specify the number of features $K$ we want to select.
2. Compute the F-statistics between $X_i$ and $y$.
3. Rank the F-statistics and select the best feature which has the highest F-statistics.
4. While (number of selected features $< K$):
   (a) Calculate correlation between selected features and unselected features.
   (b) Calculate $f^{FCQ}$ by using Equation 2.
   (c) Add the variable, which has the highest $f^{FCQ}$ score.

We applied the Maximum Relevance Minimum Redundancy (MRMR) feature selection method to the data and compared the results obtained from these variables with those from the MD data. Before applying the MRMR method, it is necessary to specify how many features will be selected. Therefore, K = 1 to 17 were first defined and the MRMR method for each K was applied to the data. AUC values from each feature were recorded and plotted as shown in SM Fig. 2. Finally, the features selected according to this method were FPG STD, HbA1c Mean, g1diabed STD, and NPHL Insulin Mean variables.

#### Performance Measures

The ACCORD dataset consists of various independent variables like continuous and categorical, and the response variables consist of binary categorical variables. Hence, Sensitivity, (or Recall), Specificity, Positive Predictive Values (PPV), (or Precision), Negative Predictive Values (NPV), Accuracy and F1 Score (harmonic mean of sensitivity and precision) are selected below to evaluate the performance of the models.

$$\text{Specificity} = \frac{TN}{TN + FP} \tag{3}$$

$$\text{PPV} = \frac{TP}{TP + FP} \tag{4}$$

$$\text{NPV} = \frac{TN}{TN + FN} \tag{5}$$

$$\text{Sensitivity} = \frac{TP}{TP + FN} \tag{6}$$

$$\text{Accuracy} = \frac{TP + TN}{TP + TN + FP + FN} \tag{7}$$

$$\text{F1-score} = \frac{2 \times \text{PPV} \times \text{Sensitivity}}{\text{PPV} + \text{Sensitivity}} \tag{8}$$

Equations 3–8 denote the number of correctly categorized positive labels by TP (the true positive) and the number of correctly classified negative labels by TN (the genuine negative). The number of labels expected to be negative but actually positive predicted is indicated by FP (false positive), while FN (false negative) indicates labels that are predicted negatively but are actually expected to be positive.
